# A complex interventions approach to delayed diagnosis in bipolar disorder. a co-produced systematic review and narrative synthesis of associated factors

**DOI:** 10.1101/2025.10.13.25337871

**Authors:** Tania Gergel, Mariam Al-Janabi, Shivangi Talwar, Haleemah Ahmed, Talen Wright

## Abstract

**Purpose:** There are widespread concerns about adverse consequences of persistent five-to-ten-year delays in diagnosing and treating bipolar disorder. Previous evidence synthesis focuses primarily on quantifying scale. This systematic review aimed to identify the complex and broad precursors, outcomes, and suggested pathways to improve diagnostic delay.

**Method:** A complex intervention approach, using co-produced narrative synthesis of quantitative and qualitative studies, is used to identify associated precursors, outcomes, and suggested avenues for improvement.

**Results:** A total of 49 generally high-quality studies from high or middle-income countries were included. This review reinforces certain findings from previous reviews regarding scale of delayed diagnosis and key associated precursors, including: history of depression, suicide attempts, earlier onset, female sex, BD-II presentation. However, we also identify a wider range of associated factors including: mixed, hypomanic, ‘atypical’, and rapid cycling presentations; suicidality in bipolar and interrelationship with diagnosis, acknowledging persistence of suicide risk following diagnosis; interrelationship with diverse mental and physical health conditions; attitudes and management of bipolar; diagnostic process in lower-and-middle income countries; clinical pathways associated with diagnosis and the complex interplay of socio-economic, cultural, and demographic factors. Suggested avenues for improvement were diverse, but highlighted screening, and the need for increased collaboration and broader research.

**Conclusions:** Our findings suggest that future research and interventions should examine delayed diagnosis in terms of its breadth and complex interrelationships with aspects of bipolar disorder which remain insufficiently understood. We also highlight the need for deeper insight into stakeholder experiences and attitudes toward bipolar management and diagnosis, particularly through qualitative, mixed-methods, and co-produced research.

## 1 Introduction

Bipolar disorder (BD) is a severe and chronic episodic mood disorder, estimated to affect 2-4% of the global population, with a high disease burden including premature mortality, high rates of suicide deaths and attempts, and poor general health and socio-economic outcomes.(1–6) Delayed diagnosis (DD) and concomitant delays accessing appropriate treatment are consistently identified by all stakeholders as key challenges for people living with BD.(1, 5, 7–16) Delays are widely acknowledged to average approximately five to ten years, with depression a common prior diagnosis and high rates of unrecognised BD amongst people diagnosed with depressive disorders.(1, 14, 15, 17) This is supported by evidence from five recent meta-analyses which provide reasonable consensus, although they identify significant between-sample heterogeneity as a limitation (7, 8, 16, 18, 19). Nevertheless, despite increasing research examining the scale of delays and some associated factors, the absence of evidence of any substantial reduction in delays suggests that our understanding of the causes, consequences, and pathways to improving BD diagnosis remains limited (8, 17, 20).

### 1.1 Applying a complex intervention framework to diagnostic delay

Within medicine, it is increasingly recognised that both diagnosis and interventions designed to reduce diagnostic delays should be viewed as complex and multifactorial interventions. The diagnostic process should be understood as a “complex, patient-centred, collaborative activity” (21), and research should use diverse approaches including qualitative work to capture complexity, interpersonal, and socio-cultural dimensions.(22–26) The value of stakeholder involvement in improving diagnostic processes and acceptability is also emphasised both in general medicine and psychiatry (22, 27), as are the particular complexities, challenges, and limitations associated with diagnosing mental disorder (28, 29). Nevertheless, a complex interventions approach has not yet been applied to research on DD and closely related phenomena, such as unrecognised BD amongst people diagnosed with depressive disorders.

### 1.2 Summary of prior research

To date, DD research and systematic synthesis has focused on synthesising epidemiological evidence to establish average duration of delayed diagnosis and treatment or rates of unrecognised BD and diagnostic conversion amongst those diagnosed with depression.

One short 2012 narrative review describes some putative adverse clinical, functional, and economic consequences of DD in BD (17), while Ratheesh et al. 2017 includes an informal summary of qualitative date relating to predictors of diagnostic conversion,(18) and Scott et al. 2022 and Keramatian et al. 2025 include informal supplementary narrative reviews of putative predictors of duration of DD, and associated lifetime outcomes and clinical characteristics respectively.(8, 16). A few qualitative studies also identify themes relating to DD.

To our knowledge, there has been: no systematic narrative synthesis of DD; no systematic synthesis including relevant qualitative literature; no co-produced research specifically targeting DD, despite DD being a priority area for people with BD.

### 1.3 Study aims

Our aim, therefore, is to identify the complex and broad precursors, outcomes, and suggested means to improve diagnostic delay, by shifting the focus of DD evidence synthesis from quantitative analysis of average duration or rates to a complex interventions approach, designed to capture breadth and complexity. We seek to identify a broader range of factors relating to DD, thereby increasing understanding, while identifying gaps and potential novel directions for progress. We use systematic narrative synthesis to integrate diverse study types, and identify diverse themes stratified into associated precursors, outcomes, and avenues for improvement.(30) Furthermore, we include qualitative studies and use co-production to prioritise incorporation of stakeholder perspectives capture interpersonal and collaborative aspects of diagnosis. This review was conceived and supervised by a researcher with lived experience of DD in BD and includes other authors with lived experience of mood disorder, as part of a partnership between Bipolar UK and University College London. This partnership aims to address stakeholder priorities identified within co-produced initiatives such as the user-led Bipolar UK Bipolar Commission, which reported average DD of 9.5 years,(31) and the James Lind Alliance Priority Setting Partnership, which identified questions relating to DD as a top-ten priority for research into BD.(10)

## 2 Methods

Our review followed Preferred Reporting Items for Systematic Reviews and Meta-Analyses guidelines, and is registered on PROSPERO, CRD42024537132 (see Supplementary Information for further details).

### 2.1 Search strategy, inclusion and exclusion criteria

We searched MEDLINE, PsycINFO, Embase, and Web of Science from database inception until 15^th^ July 2024, supplemented by reference searches of included articles and relevant reviews until January 2025. Key search terms included the main constructs and their derivatives, designed to capture core concepts of delay (including failure to recognise BD and instances where diagnostic conversion from depression to BD is indicated), diagnosis and/or treatment, and BD (for full search strategy see Supplementary Information). Eligible studies comprised quantitative, qualitative, or mixed-methods studies published in English in peer-reviewed journals with adults diagnosed and/or treated for BD as a primary population, and with delayed diagnosis and/or treatment of BD identified as primary or secondary exposure or theme/subtheme. We included studies with diagnostic conversion from depression to BD or unrecognised BD amongst people with a depression diagnosis as a primary exposure, viewing such studies as highly relevant to DD, given that evidence identifies depression as the most common precursor diagnosis to BD diagnosis. We did not assess grey literature sources or studies focused on paediatric bipolar with populations primarily comprising participants ≤ 18 years, given our primary focus on adult-pattern BD.(19)

### 2.2 Study selection

Titles and abstracts were extracted into endnote for deduplication. TG and MA independently screened titles, abstracts, and full texts. At each stage, disagreements were resolved by discussion between the two raters, with input from TW, HA, and ST in complex cases. Studies with ≤5 participants and studies assessed as below moderate standards were excluded.

### 2.3 Data extraction

A data extraction was created in Excel including study location; methods (including evidence of co-production); sample size and characteristics; primary diagnostic delay exposure and/or focus of study; average DD length; and main study findings relating to diagnostic and treatment delays. Quantitative duration data was classified following latencies used in previous meta-analysis (see Table 2), with supplementary descriptions of any variance. Data for all studies was extracted independently by MA and TG. Disagreements were resolved by initial discussion between the two reviewers, with all authors helping to resolve any uncertainties or ongoing disagreements and carrying out final checks.

### 2.4 Quality assessment

The quality of included studies was assessed using the Kmet et al. Standard Quality Assessment Criteria for evaluating primary research papers from a variety of fields (32). The Standard Quality Assessment Criteria were selected as having been validated and used in previous systematic reviews including both quantitative and qualitative designs. Quality of included studies was assessed by one reviewer (MA), with a second reviewer (TG) independently conducting quality assessment of a randomly generated sample of 25% of studies. There were high levels of agreement between reviewers and any conflicts in quality appraisal were discussed with the wider review team until consensus was reached. All studies were attributed equal value in terms of contributing to the summary findings.

### 2.5 Data synthesis

We analysed findings using qualitative narrative synthesis, a methodology suited to capturing breadth and complexity, and to managing the substantial heterogeneity of focus, methodology, variables, measurements, and study populations in the relevant literature.(30) All stages followed an integrated approach, with data extraction, analysis, and presentation of findings executed within a unified framework, regardless of study type. The approach was qualitising, insofar as: identification of themes was not based purely on statistical significance or frequency of findings; and analysis was thematic, allowing a small quantification element in noting frequency of appearance of themes.(30) We used extracted data to identify themes and subthemes for associated precursors, outcomes, and putative avenues for improvement. We used an iterative deductive methodology combining guidelines for narrative synthesis in systematic reviews and thematic analysis (for more details see Supplementary Information 7).(33, 34) TG and MA coded all articles independently using Microsoft Excel, returning to full texts of for any clarification and expansion of findings. An inductive approach was then used to refine and expand initial coding and themes through an iterative process until all articles were analysed, with codes and themes reviewed regularly by TW and ST, until reaching consensus.

## 3 Results

Our search identified 8334 studies after deduplication and screening by article type, with 7843 excluded by title. 362 articles were excluded after screening abstracts. Of 129 full-text records screened, 40 met inclusion criteria. After reference searches and full text screening of 14, nice further studies were added. This totalled 49 studies included in the systematic review (see Figure 1).

**Figure 1:**
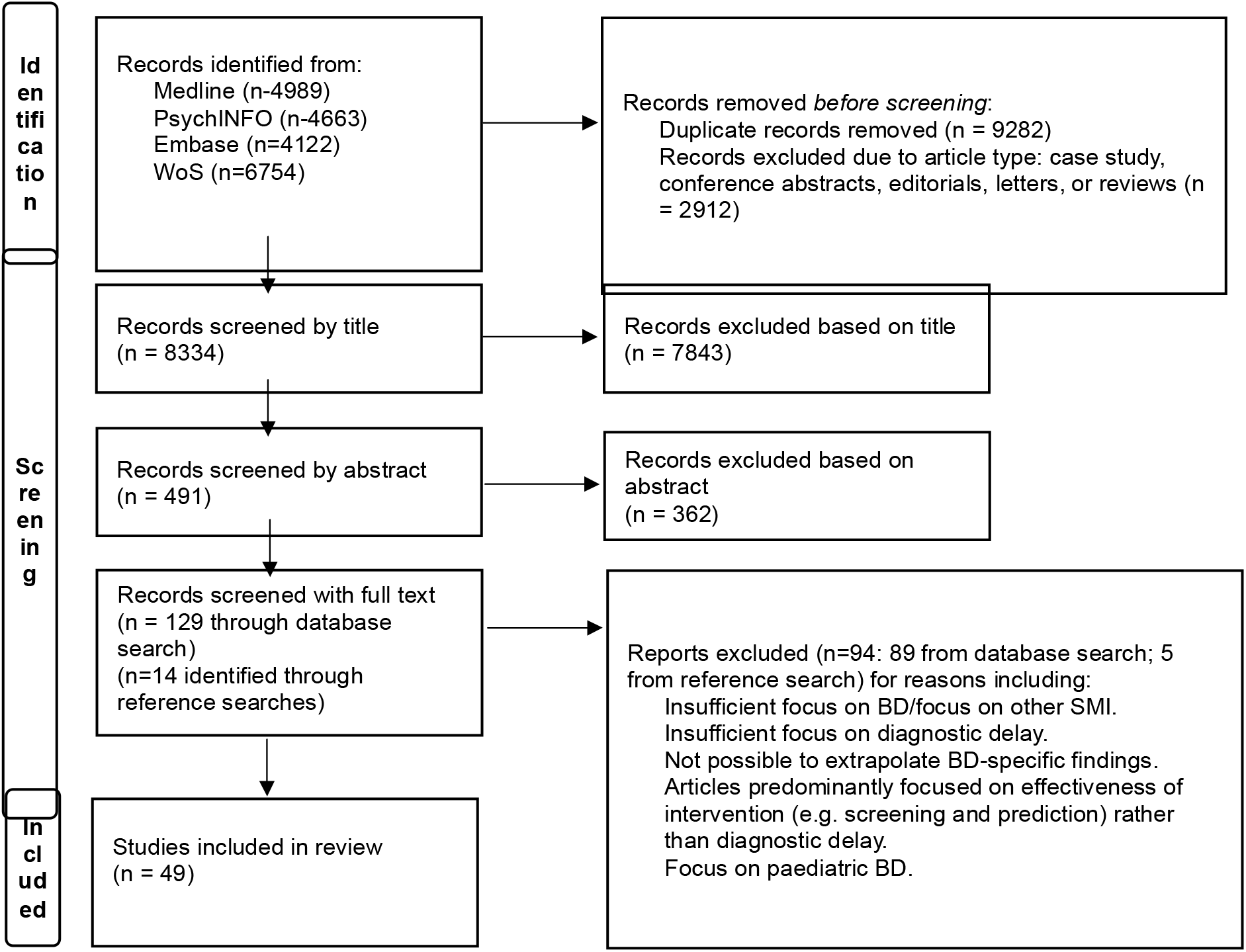
Study Flow Diagram.

### 3.1 Study characteristics

Study characteristics are presented in Table 2.

Studies were conducted in 27 countries, with 22 in Europe, five in North America, four in Australasia, 11 in Asia, five in South America, and two in Africa. All studies took place in high or middle-income countries, with two middle-income based studies exploring the interrelationship of socio-economic factors and DUB (duration of untreated bipolar disorder) in developing nations.(35, 36) 39 studies were observational epidemiological studies, with DD, DUB, or unrecognised BD and diagnostic conversion from depression to BD as primary or secondary exposures. The primary exposure was delayed diagnosis or treatment in 13 studies, associated precursors or outcomes in 14 studies, and rates of diagnostic conversion to BD or unrecognised BD amongst people diagnosed with depressive disorders in 12 studies. We also included ten qualitative studies which identified DD as a theme/subtheme. Involvement of people with lived experience of BD in design, implementation, or dissemination of research was reported in three qualitative studies,(37–39) but in no epidemiological studies.

There was substantial variation in the primary focus of included studies. 12 epidemiological studies focused broadly on clinical and/or socio-demographic associated precursors or outcomes. Other studies focused on specific associations or populations: anti-depressant use;(40–42) clinical cost of DUB;(43) relapse frequency;(44) manic or depressive polarities;(35, 45) increased suicidality;(46–48) psychotic symptoms;(49) cannabis use;(50) general medical comorbidities.(51) Some comparative studies examined whether DD duration is reducing;(52, 53) variation between BD-I (Bipolar I Disorder) and BD-II (Bipolar II Disorder), (48, 54) duration in LMICs (lower-and-middle-income countries);(36) variation between early onset and later onset BD.(55, 56) Nine qualitative studies focused on lived experience of BD examining challenges;(37, 57) healthcare;(58, 59) diagnosis;(38, 59–62); suicide;(39) research priorities.(63) One qualitative study explored clinical BD research priorities and correlation with service user priorities.(11)

### 3.2 Quality assessment

We judged 28 of the studies to be of high methodological quality (with 22 studies judged high and 16 very high), with the remaining studies medium (N=9) and low (N=2) [See Quality Assessment Table in Supplementary information].

### 3.3 Qualitative synthesis of scale of delay and unrecognised BD

All studies measuring delay duration reported long delays in diagnosing and treating BD and high rates of unrecognised BD, despite heterogeneity of study methods, measurements, latencies, and populations. The majority of studies (17/25) presenting average delays in diagnosis or treatment found averages ranging between 5.16 and 10.4 years. Nine of 12 (75%) studies presenting unrecognised BD rates found averages ranging between 18.21% and 39.84%. Further details about these findings, including population-based factors for studies with unusually high or low averages can be found in Supplementary information.

### 3.4 Narrative synthesis of precursors, consequences, and avenues for improving delays in diagnosing and treating BD

#### 3.4.1 Precursors associated with delays in diagnosing and treating BD

We identified five precursor themes, with four involving clinical factors, including illness presentation, course, and management. A positive association with increased delay was identified for BD subtypes associated with mixed, hypomanic, or depressive presentations, and with prior depression diagnosis. Studies examining associations with history of anxiety disorders, ADHD (Attention Deficit Hyperactivity Disorder), PTSD (Post-Traumatic Stress Disorder), personality disorders, multiple prior diagnoses, and general medical conditions also found positive associations with increased delay duration. However, results were more mixed for substance abuse and psychosis. For substance abuse, findings were a mixture of positive, negative, or no association. While history of psychosis was generally associated with shorter delays, some studies identified psychotic disorder diagnosis as a common precursor to BD diagnosis, and one qualitative study linked this phenomenon to predominantly hypo/manic presentations.

For illness course, multiple epidemiological studies found earlier age of onset positively associated with increased delay duration, while 12 of 15 studies examining associations with suicide attempt history showed positive associations, and one study showed positive associations with reduced interepisodic recovery. Findings relating to family history of BD were varied and inconclusive.

Multiple studies showed diverse positive associations between elements of clinical pathways and longer delay duration. These included particular diagnostic frameworks, poor clinical knowledge, primary care, shorter treatment periods, and increased frequency of medication changes. However, findings relating to history and pattern of hospitalisations were mixed. Three clinical pathway subthemes were identified in qualitative studies as factors potentially associated with increased DD, with three studies highlighting restricted knowledge and treatment options within primary care, (37, 58, 59) one identifying clinical non-disclosure of diagnosis,(61) and four studies suggesting that particular mood states may not be witnessed by a clinician.(37, 38, 59, 60)

Finally, positive or putative associations with various demographic, socioeconomic and cultural factors, and increased duration of DD/DUB were presented. These included poor psychoeducation, stigma, and concerns surrounding BD, lower socio-economic status, and female sex, although one study on diagnostic conversion found associations with male sex (64) and one study found higher functioning in BD-I associated with longer DD.(65) Findings were inconclusive in relation to social isolation and duration of DD/DUB. Additional factors and complexities reported in qualitative studies included clinical reluctance to diagnose BD due to stigma, high-functioning presentations, or concerns about overdiagnosis; how clinical failure to diagnose BD may result in inaccurate suicide risk assessment; clinical prioritisation of physical healthcare issues in primary care; service user/family failure to recognise certain mood states as needing medical attention and reluctance to report hypomanic/manic symptoms or accept diagnosis, although non-acceptance of diagnosis did not necessarily cause rejection of BD-specific treatment.

#### 3.4.2 Outcomes associated with delays in diagnosing and treating BD

We identified three themes amongst outcomes associated with delays, all pointing to association of longer delays with poorer outcomes post-diagnosis and treatment. Poorer illness course outcomes were: worsened overall illness course, increased incidence of suicide attempts, poorer outcomes following anti-depressant monotherapy, worsened physical health, and increased incidence of comorbidity with anxiety disorder and substance abuse. Poorer clinical outcomes related to increased frequency of hospitalisations, cost of care, and delays accessing treatment. Qualitative studies also suggested associations with decreased service user faith in the healthcare system and insufficient information at diagnosis resulting in longer delays in accessing treatment. Finally, increased delay duration was positively associated with multiple poorer socio-environmental outcomes.

#### 3.4.3 Suggested avenues for improving BD diagnosis

We identified four themes amongst avenues for improving the diagnostic process amongst included studies, with three themes relating to potential interventions and one relating to research. Two themes related to aspects of clinical care, the need to implement some form of routine and improved screening process and the need for improved services and clinical education. Screening for BD was a very common suggestion, particularly amongst service users presenting with depression or suicide attempts. There was a broad variety of subthemes, focusing on looking for particular BD subtypes amongst particular cohorts or family history. Suggestions for improving services and clinical education included changing clinical pathways and education, improving resources, and introducing BD early intervention services. Our third theme was identified within multiple studies, suggesting that community and stakeholder psychoeducation could improve diagnosis and reduce stigma. Suggestions for improving research ranged from general suggestions about DD research to targeting specific areas, such as differentiating BD subtypes or BD depression from major depressive disorder and research on diagnostic processes and assessment.

Stakeholders in qualitative studies highlighted the potential usefulness of collaborative measures and interpersonal exploration, including team-based assessment with family involvement; symptom tracking; expanded psychoeducation, involvement, and co-management; increased clinical disclosure; collaborative exploration of diagnostic experience and implications to facilitate stakeholder acceptance; qualitative work exploring clinical assumptions surrounding high-functioning presentations.

## 4 Discussion

### 4.1 Summary of findings in comparison with prior research

We identified diverse and multifactorial associated precursors, outcomes, and suggested avenues for improvement, which support stakeholder recognition of widespread five-to-ten-year averages for DD, associated negative outcomes, and urgent need for timely BD diagnosis and appropriate treatment.(1, 5, 10, 11, 14, 31, 63, 66–71) Consistent with prior systematic reviews, we found no evidence of substantive delay reduction despite longstanding recognition of this issue and extensive epidemiological research.(7, 8, 16)

Our findings were broadly consistent with previous reviews in identifying some widely reported precursors of longer DD, including: history of depressive states; predominance of depressive polarity or BD-II sub-type; earlier age of onset; female sex; history of suicide attempts (see Table 1). This underscores the importance of prioritising improved detection amongst individuals meeting these criteria. However, our approach also facilitated identification of a broader range of DD-related factors, which expand upon and sometimes challenge previous findings.

**Table 1.** Summary of results from previous systematic reviews and meta-analyses. Abbreviations DD = Duration of delay between onset of illness and diagnosis. DUB = Duration of untreated bipolar disorder (delay between onset or seeking professional help and clinical management with appropriate treatment). DC = Rates of diagnostic conversion in sample from major depressive disorder / unipolar depression diagnosis to BD diagnosis. UB = Unrecognised bipolar disorder amongst those with a diagnosis of depression. DHS = Duration of delay between onset of illness and seeking clinical help. DUDB = Duration of delay between onset of illness and clinical management with appropriate treatment or BD diagnosis.

| Authors / date | No. of studies included | Primary exposure/s and findings | Associations reported. | Qualitative synthesis included | Main qualitative synthesis results noted |
| --- | --- | --- | --- | --- | --- |
| Dagani et al. 2017 | 27 | Mean DUB: 5.8 years. |  |  |  |
| Ratheesh et al. 2017 | 56 (qualitative synthesis of rates); 35 (pooled quantitative analysis); 24 (qualitative or quantitative analysis of predictors). | Mean DC (with 12-18 years mean follow-up): 22.5% of adults and adolescents. | **DC Predictors:**<br>Family history of BD.<br>Lower age depressive onset.<br>Psychotic symptoms. | Informal summary of qualitative data relating to predictors, with results relating to phenomenology and course of depression; subthreshold (hypo)manic features; comorbidity; family history and childhood risk factors. However, there is no systematic synthesis. | DC predictors:<br>Subthreshold manic symptom |
| Daveney et al. 2019 | 10 | Mean UB amongst patients in primary care: 17% |  |  |  |
| Scott et al. 2022 | 59 | Mean DD: 6.7 years<br>Mean DUB: 5.9 years.<br>Mean DHS: 3.5 | **Longer delay precursors:**<br>Exclusively outpatient treatment.<br>Early age of onset.<br>Female sex (especially BD-II). | Informal supplementary narrative review including simple schematic to demonstrate variables most commonly identified as putative predictors of longer or shorter DD and perceived magnitude of effect. | Predictors of longer DD: history of childhood neglect and/or suicide attempts, depressive onset, BD-II, female sex, and early age at onset. Predictors of shorter DD: psychotic onset. Uncertainties relating to substance abuse and family history of |
|  |  | years. | **Shorter delay precursors:**<br>Psychotic onset.<br>Residing outside N. America<br>Access to early intervention services.<br>Older participant age. |  | BD as predictors. |
| Keramatian et al. 2025 | 23 | Mean DUBD: 9.1 years. | **Longer delay:**<br>Early age of onset.<br>Family history of BD.<br>Comorbid anxiety or substance abuse.<br>Lifetime suicide attempts.<br>**Shorter delay:**<br>BD-I diagnosis.<br>Lifetime psychotic symptoms. | Informal supplementary narrative review of studies reporting on lifetime outcomes and clinical characteristics. | Lifetime outcomes and clinical characteristics associated with longer DUBD included: frequency of previous mood episodes, with mixed findings relating to manic episodes; inconclusive findings relating to history of rapid-cycling BD; mainly positive correlations with history of anxiety disorder; mixed findings relating to medical comorbidity; mixed findings relating to lifetime history of suicide attempts; mixed findings relating to history of psychiatric hospitalisations; either negative or no correlation with history of psychotic features; mixed findings relating to lifetime substance abuse. |

**Table 2.**
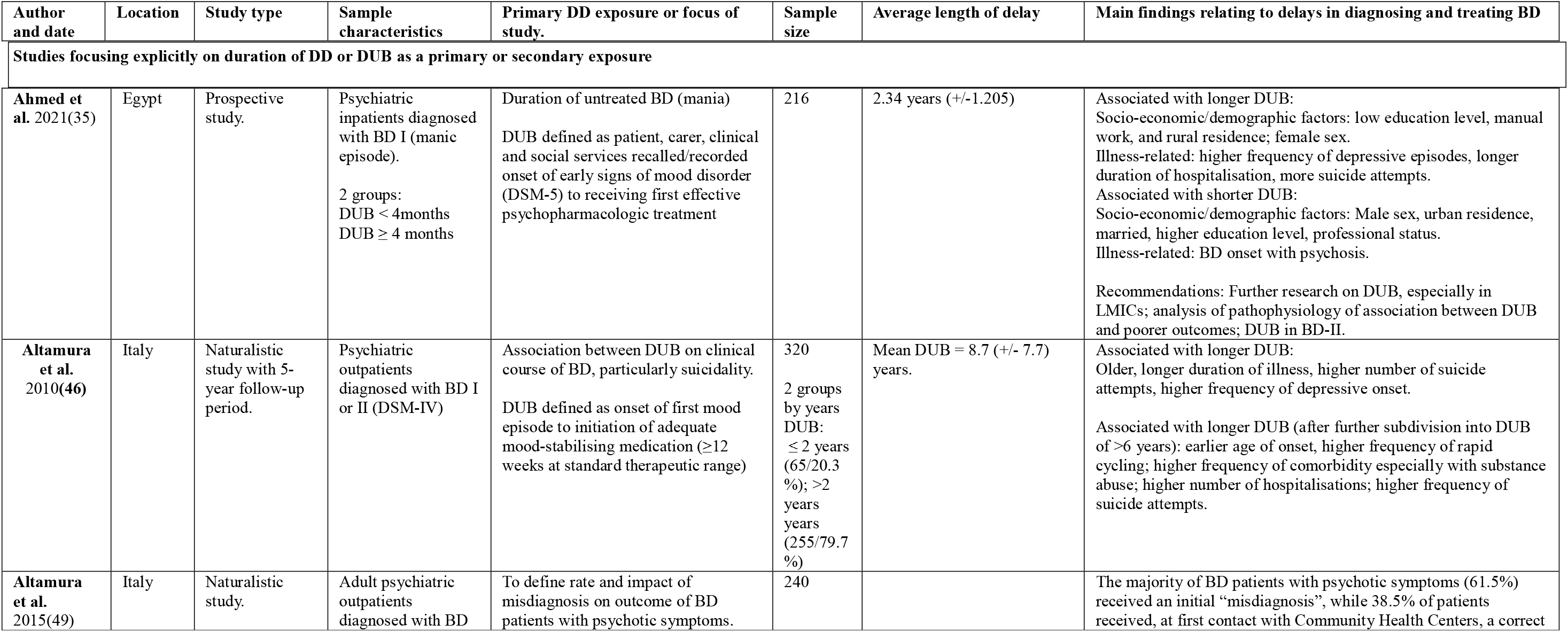
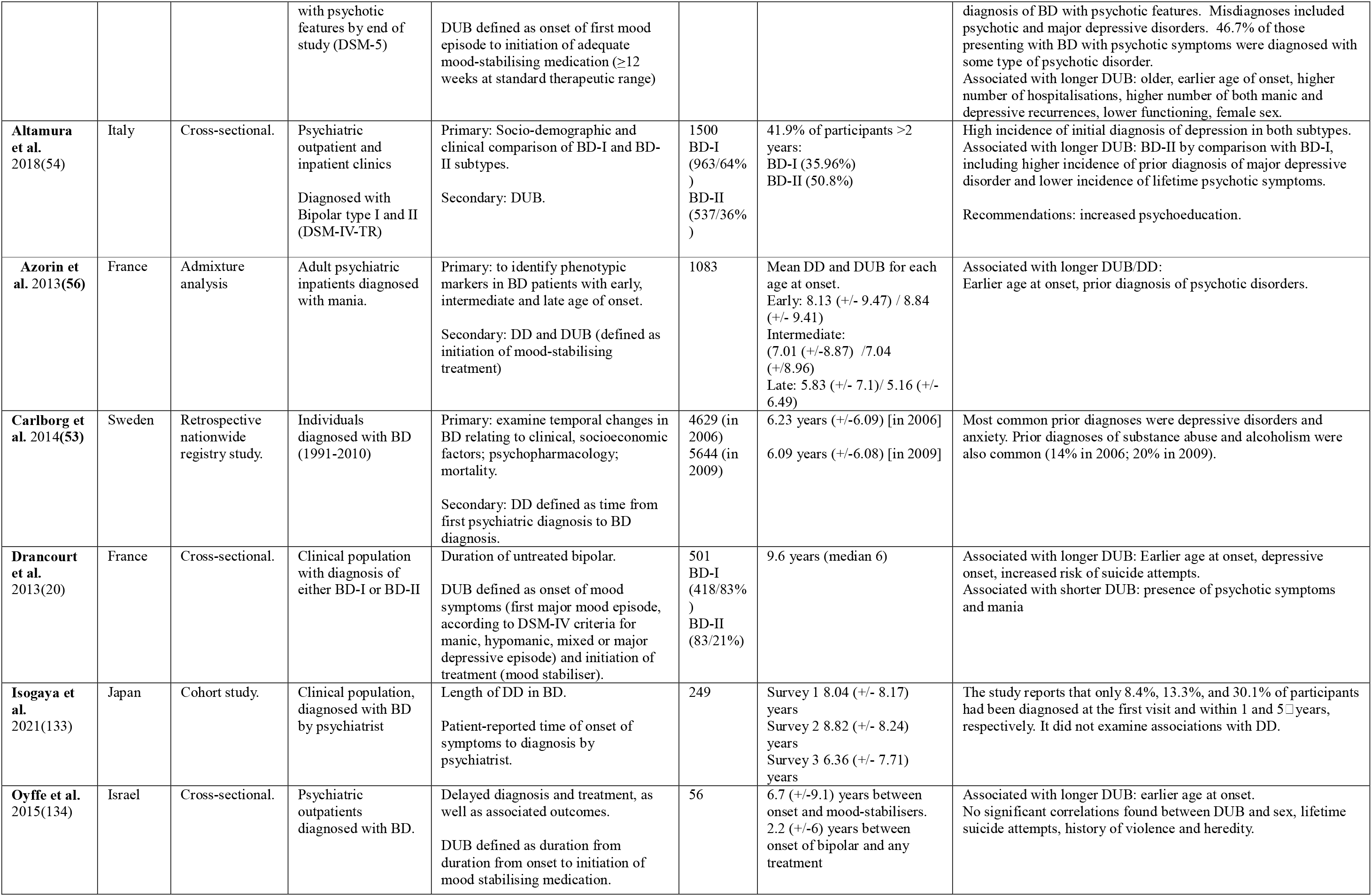
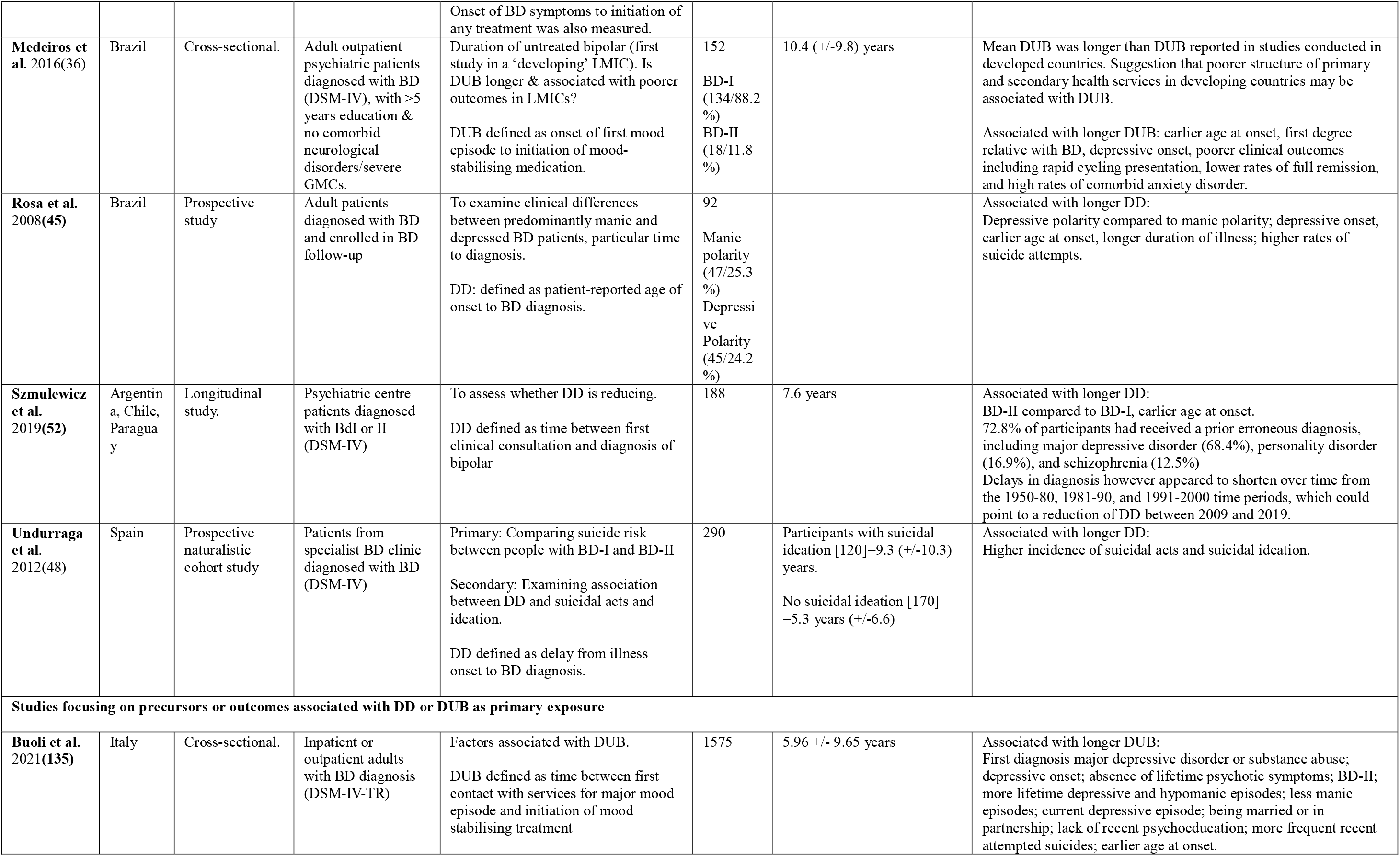
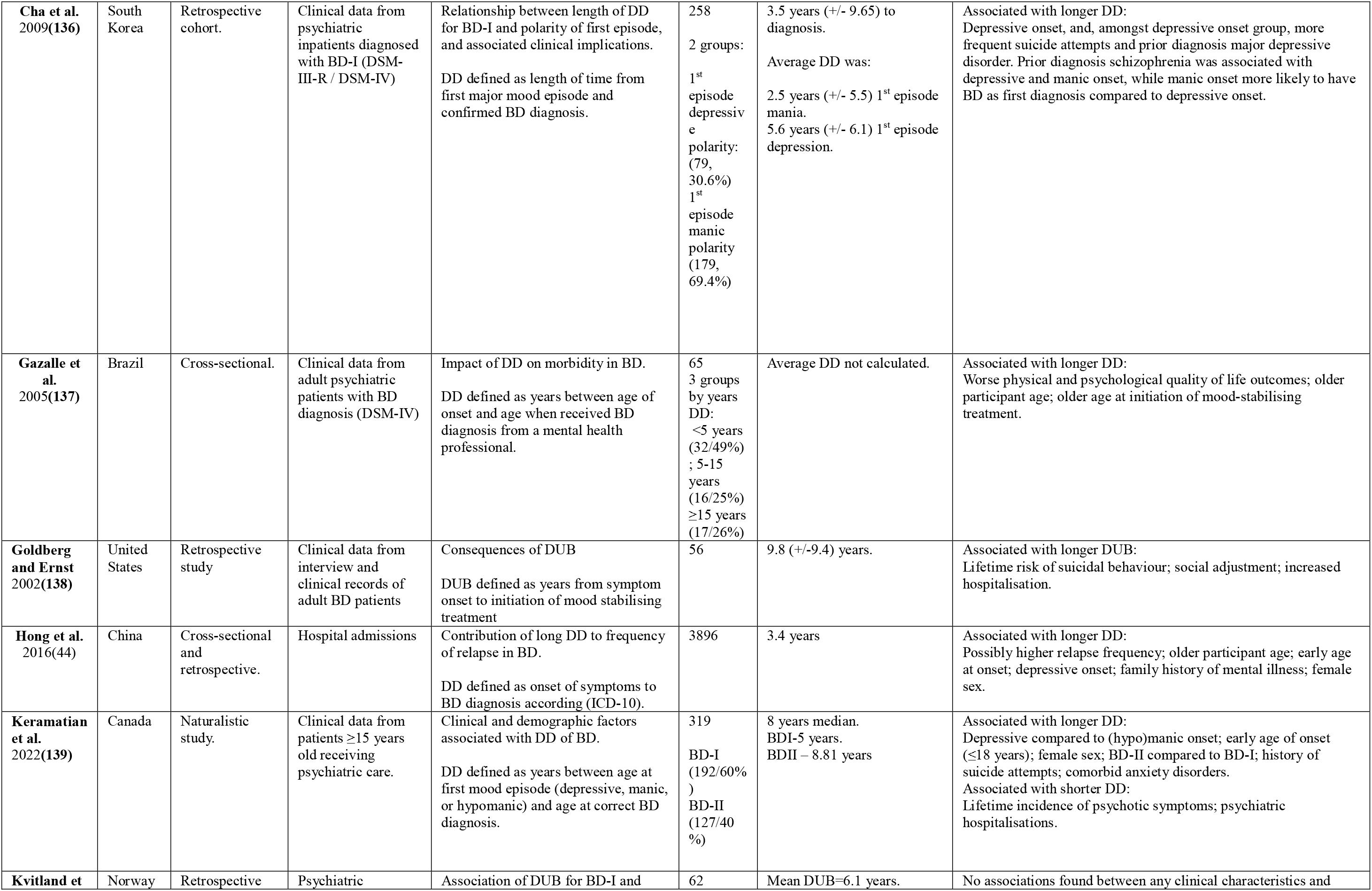
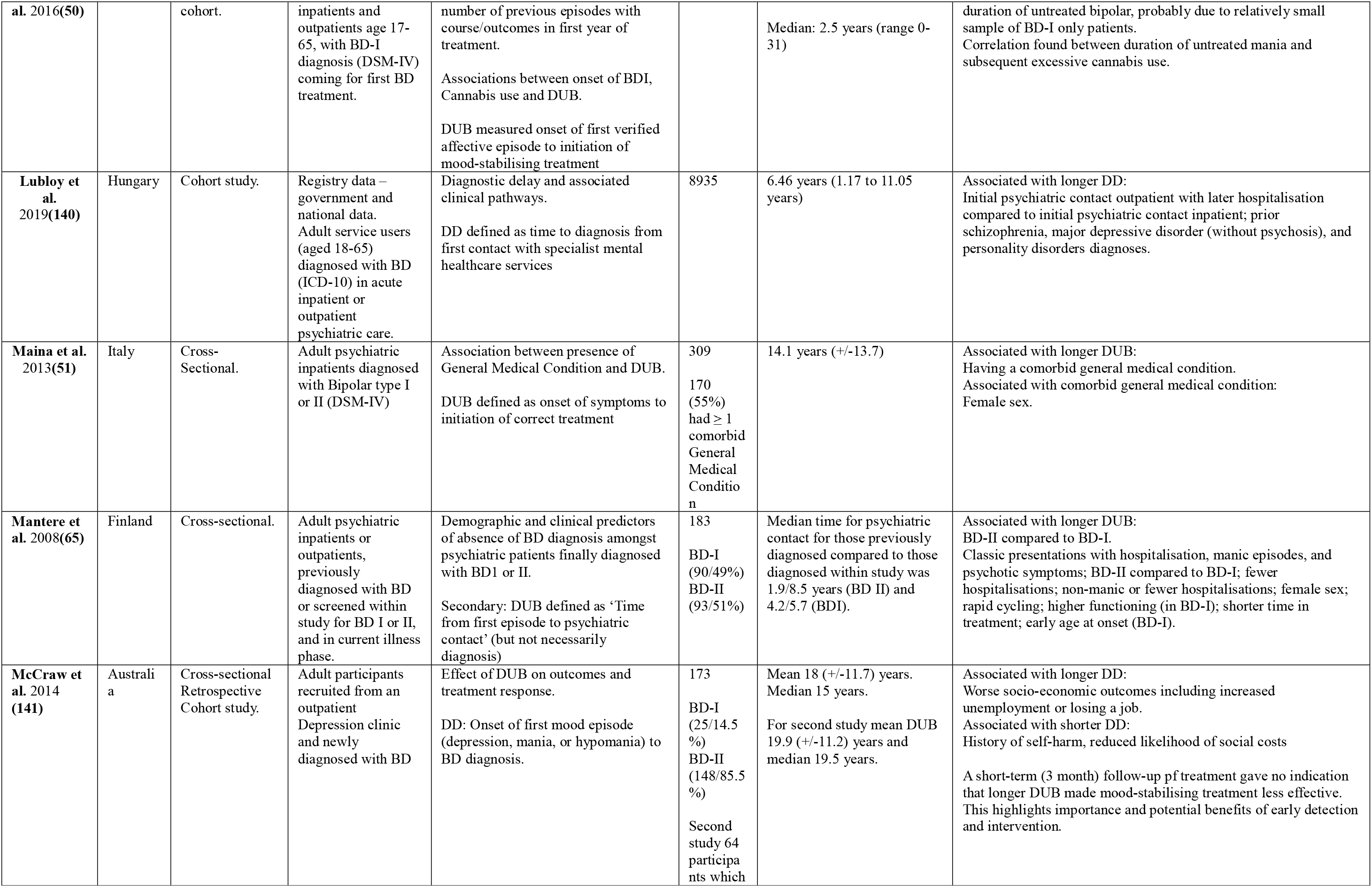
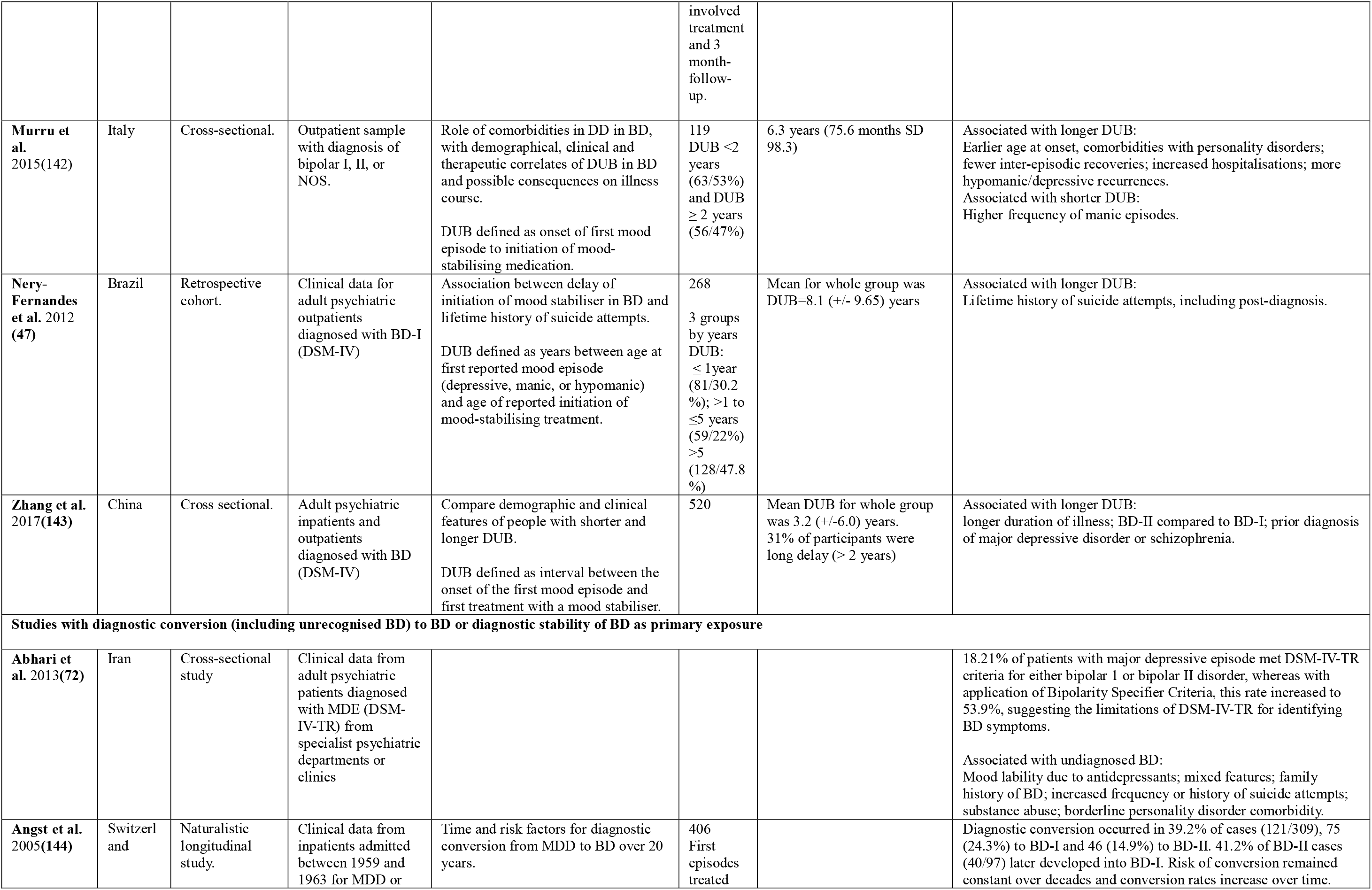
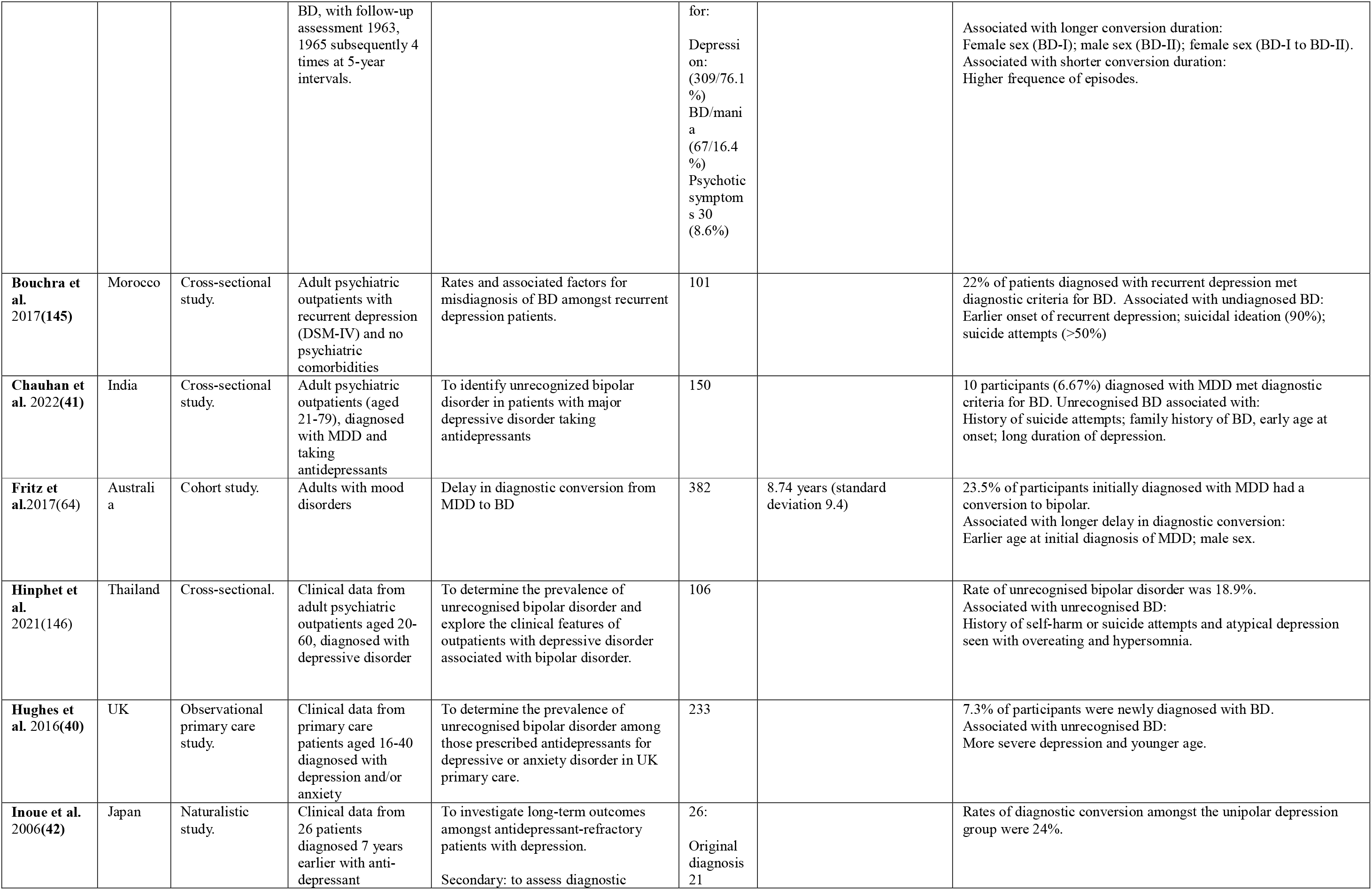
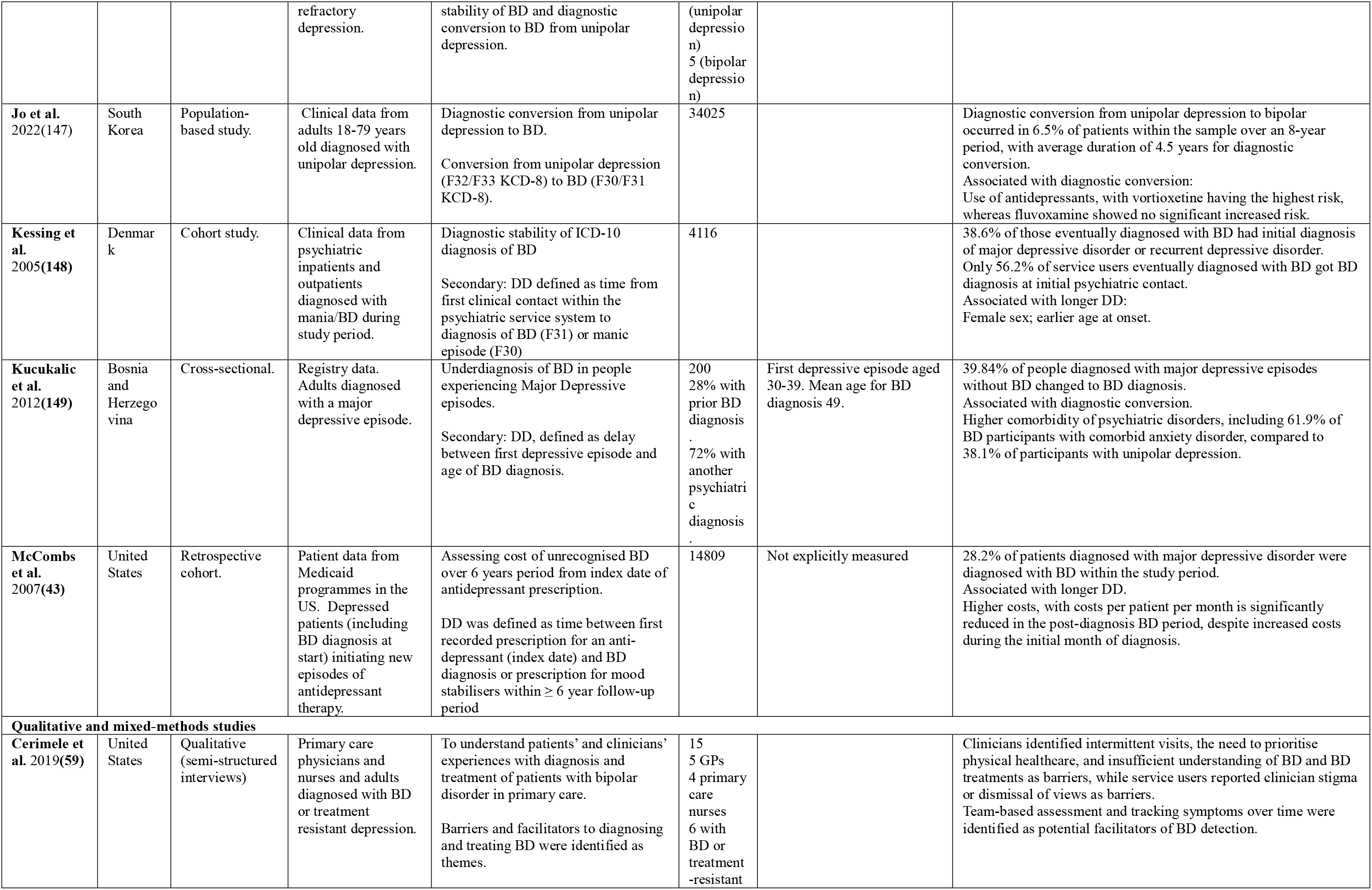
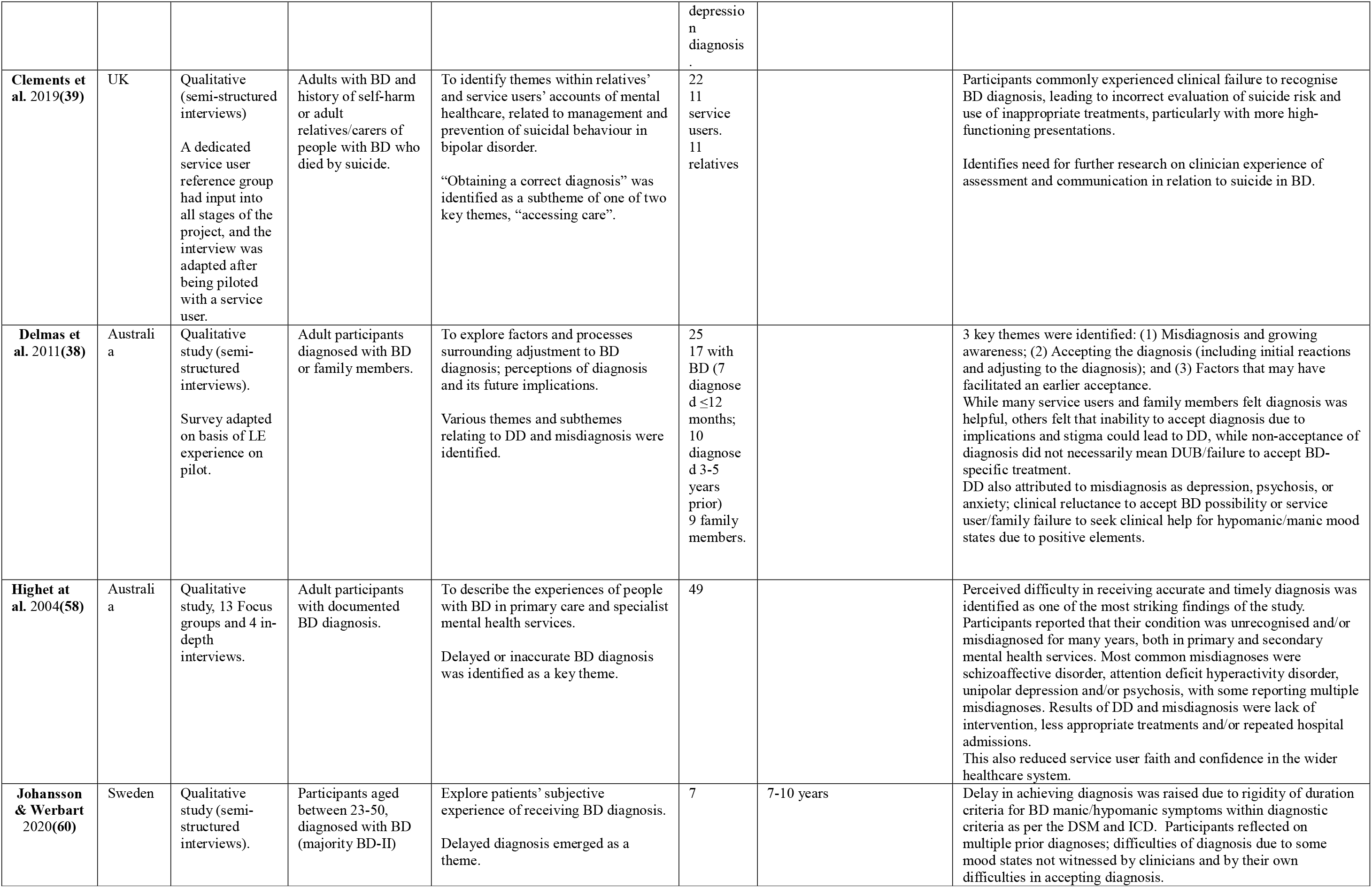
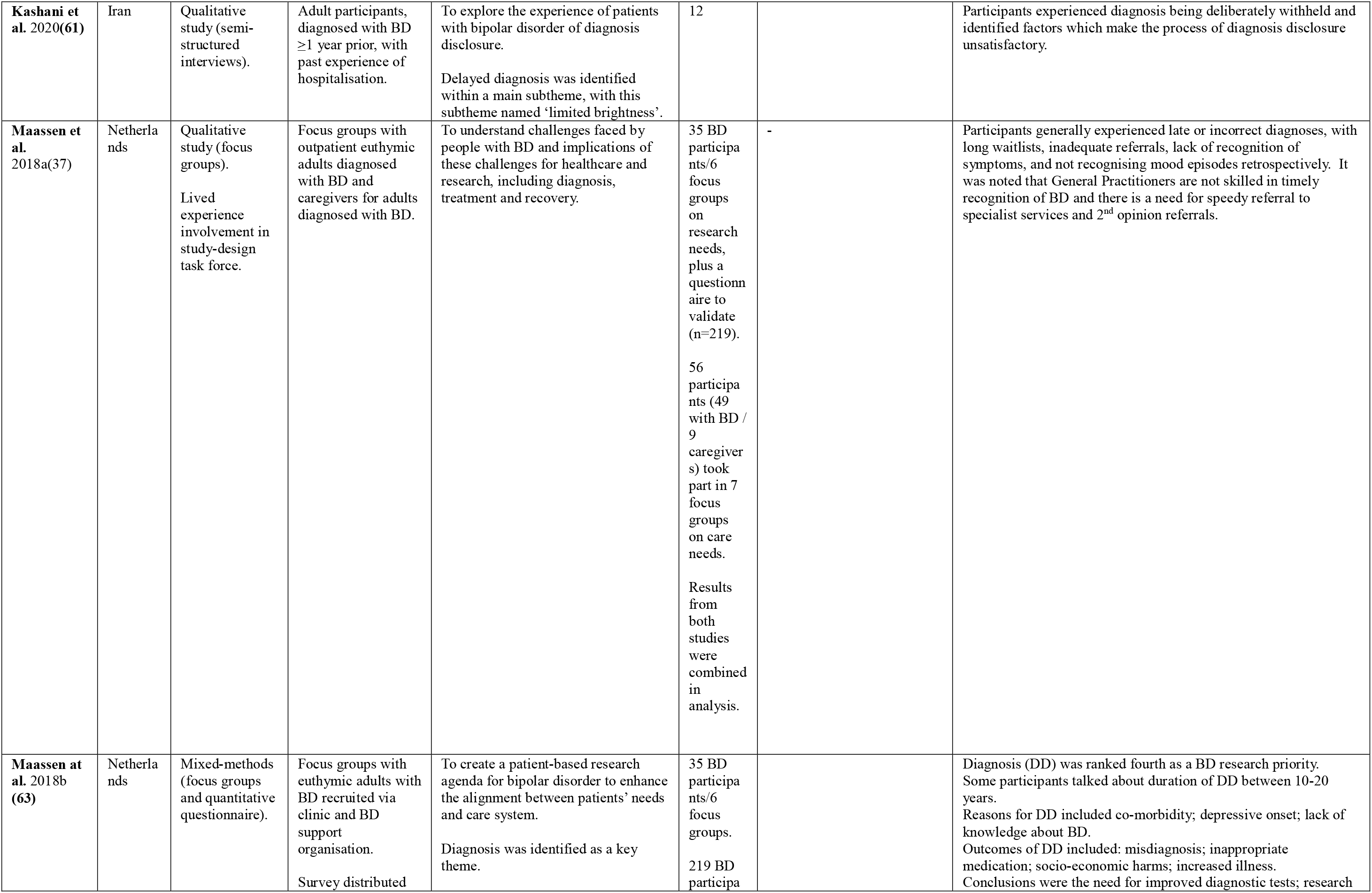
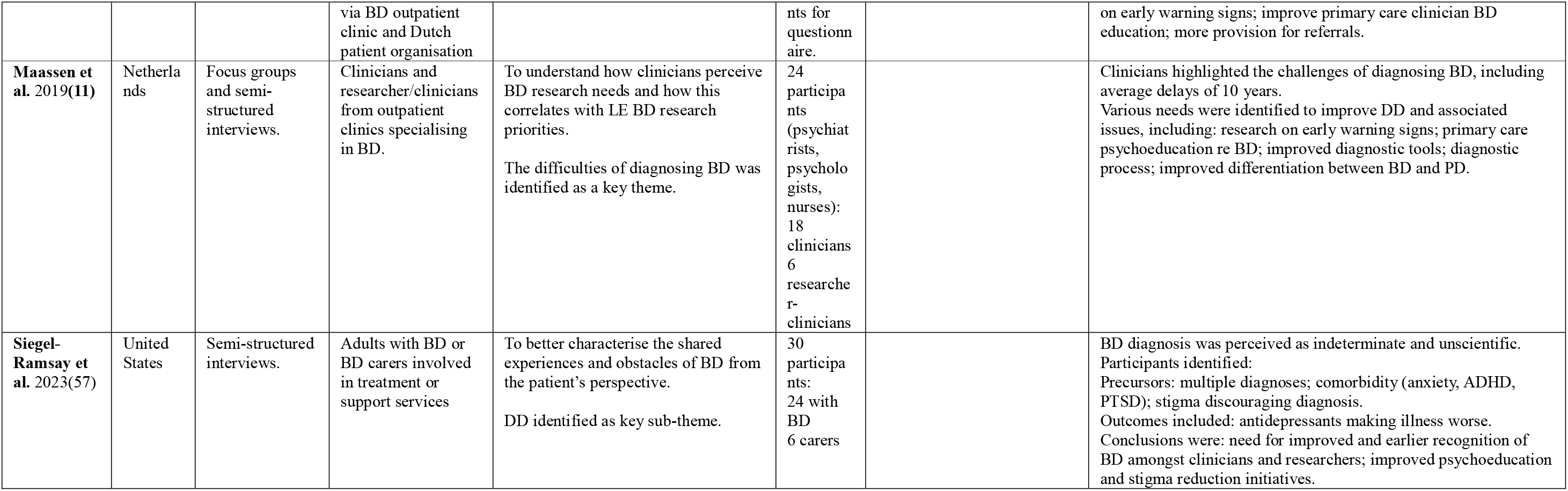
Study characteristics. Abbreviations DD = delayed diagnosis. DUB = duration of untreated bipolar disorder. DC = diagnostic conversion. BD = bipolar disorder. MDD = major depressive disorder

**Table 3.** Themes relating to associated precursors, outcomes, and suggested avenues for improvement.

| **1. Delayed diagnosis – associated precursors** |  |  |
| --- | --- | --- |
| **Sub-theme** | **Studies including subtheme (N=)** | **Summary of key findings relating to each subtheme** |
| **Theme 1: Association of delayed diagnosis with particular BD subtypes or precursors** |  |  |
| Higher frequency of mixed/hypomanic episodes/onset or atypical depression | 5(20, 72, 135, 142, 146) | A positive association was found between presenting with mixed or hypomanic features and increased delays in diagnosis and treatment. |
| Lower incidence of manic episodes/manic onset | 5(20, 45, 65, 135, 142) | Some studies found that a history of manic episodes or manic onset in BD was associated with lower delays. |
| BD-II longer DUB than BD-I | 8(20, 52, 54, 65, 135, 139, 141, 143) | Studies which compared duration of delayed treatment or diagnosis between patients receiving a diagnosis of BD-I or BD-II found that BD-II was associated with longer delays. |
| Rapid cycling | 2(20, 65) | A positive association was found between a rapid-cycling BD presentation and increased delays in diagnosis and treatment compared to other BD subtypes. |
| **Theme 2: Association of delayed diagnosis with a history and/or prior diagnosis or comorbidity with other health conditions** |  |  |
| Depression | 28(20, 35, 36, 38, 40-42, 45-47, 52-54, 58, 63-65, 135, 136, 139, 141-146, 148, 150, 151) | Over half the studies showed a strong positive association between a depressive presentation and/or depressive onset, history of major depressive episodes, major depression diagnosis, and increased duration of DD or DUB/ high rates of DC to BD. |
| Psychotic disorders | 17(20, 35, 49, 52-54, 56, 58, 65, 135, 136, 139, 141, 143, 148, 151) | Findings relating to associations with a presentation, history or prior diagnosis of psychotic disorders were mixed. It was generally found that a history of psychosis was associated with shorter DD and DUB compared to BD patients without a history or presentation of psychosis. However, a psychotic disorder diagnosis was also found to be a relatively common prior diagnosis before BD diagnosis and amongst specific BD with psychosis, a psychotic disorders misdiagnosis was very common. Prior psychosis disorder diagnosis may be more common in hypomanic/manic presentations. |
| Substance abuse disorder | 11(50, 53, 65, 72, 135, 139, 141, 142, 148, 151) | Findings relating to associations between a presentation, history or prior diagnosis of substance abuse were mixed, with half these studies showing a positive association, and half of these studies finding no association or a negative association. |
| Anxiety disorder | 7(38, 53, 57, 139, 150, 151) | A positive association was found between presentation, history, prior diagnosis or comorbidity with anxiety disorder and increased duration of DD or DUB or rates of DC to BD. Prior anxiety disorder diagnosis may be more common in hypomanic/manic presentations. |
| ADHD | 2(57, 58) | A positive association was found between presentation, history, prior diagnosis or comorbidity with ADHD and increased duration of DD or DUB or rates of DC to BD. |
| PTSD | 1(57) | A positive association was found between presentation, history, prior diagnosis or comorbidity with PTSD and increased duration of DD or DUB or rates of DC to BD. |
| Personality disorders | 4(52, 72, 142, 151) | A positive association was found between presentation, history, prior diagnosis or comorbidity with personality disorders and increased duration of DD or DUB or rates of DC to BD. |
| Multiple misdiagnoses | 4(57, 58, 60, 63, 151) | A positive association was found between a history of multiple prior diagnoses and increased duration of DD or DUB or rates of DC to BD. |
| General medical condition (comorbidity) | 2(51, 59) | A positive association was found between comorbidity with a generalised medical condition and increased duration of DD or DUB. Clinicians reported that treatment time in primary care is devoted to physical health conditions instead of BD. |
| **Theme 3: Association of delayed diagnosis with illness onset, history, and course** |  |  |
| Earlier age of onset | 15(20, 36, 41, 44, 45, 49, 52, 55, 65, 135, 139, 142, 147, 148) | A positive association was found between earlier age of BD onset and increased duration of DD or DUB or rates of DC to BD. |
| History of suicide attempts or recent suicide attempts | 15(35, 39, 45-48, 65, 134-136, 139, 141, 145, 146) | 12 studies, including three studies in which association with suicidality was a primary exposure, reported a positive association between increased history of suicide attempts and increased duration of DD or DUB or rates of DC to BD. 3 studies reported no significant association. One qualitative study identified DD as contributory factor in inaccurate risk assessment. |
| Family history of BD | 4(36, 44, 72, 139) | Findings relating to associations with a family history of BD or SMI and increased duration of delayed diagnosis or treatment were mixed, with some studies reporting positive association and some finding no significant association. |
| **Theme 4: Association of delayed diagnosis with particular elements of the clinical pathway** |  |  |
| Weakness of diagnostic frameworks, process and poor clinical education | 12(20, 35, 38, 46, 58-60, 64, 65, 72, 134, 136) | A number of studies found associations between particular diagnostic classificatory frameworks and increased DD and DUB, while other studies pointed to problems with clinical knowledge or judgement as explanatory reasons for longer duration of delayed BD diagnosis and treatment. |
| Non-disclosure of diagnosis by clinician. | 1(61) | Deliberate withholding of diagnosis from service users by clinicians. |
| Primary care | 3(37, 58, 59) | Data from two of the qualitative studies suggested associations between lack of knowledge and restricted treatment options available within primary care and increased duration of DD and DUB. |
| Hospitalisation | 5(43, 65, 139, 142, 151) | Findings relating to associations with history of hospitalisation and duration of DD and DUB or rates of DC were very mixed, with some studies finding positive associations, some finding negative associations and some pointing to very positive associations with very specific patterns, such as initial outpatient contact, with subsequent inpatient care.(151) |
| Shorter periods in treatment | 1(65) | A positive association was found between shorter prior periods of treatment and increased duration of DUB. |
| Increased frequency of medication changes | 1(142) | A positive association was found between increased frequency of medication changes and increased duration of DUB. |
| Not all mood states witnessed by clinician | 4(37, 38, 59, 60) | Given frequency and variety of mood states and differing levels of severity, not all states would be witnessed by healthcare professionals, people would not necessarily recognise certain mood states as needing medical attention, and both service users and family members might not report hypomanic/manic symptoms due to positive aspects of experience. |
| **Theme 5: Demographic, socioeconomic and environmental associations of DD** |  |  |
| Poor psychoeducation, stigma and concerns affect diagnostic process or acceptance | 9(35, 37, 38, 57, 59, 60, 63, 135, 136) | Data from some of the qualitative studies, together with putative precursors suggested within some of the epidemiological research, suggested associations between a lack of psychoeducation and awareness surrounding BD, stigma associated with BD diagnosis, clinician reluctance to diagnose BD due to stigma, overdiagnosis, and assumptions about high-functioning presentations, service user and family reluctance to accept diagnosis and increased duration of DD and possibly DUB, although failure to accept diagnosis did not necessarily lead to rejection of BD-specific treatment.. |
| Lower socio-economic status | 2(35, 65, 141) | A positive association was found between lower socioeconomic status, including employment and education status, as well as factors which may lead to reduced access to healthcare and increased duration of DD and DUB. However, higher social functioning in BD-I was also associated with longer DUB. |
| Social isolation | 3(35, 135, 141) | Findings relating to associations between social isolation and duration of DD and DUB were inconclusive, with 2 studies finding positive association between living alone and increased DD or DUB, while one found that a history of at least one long-term relationship was positively associated with increased DUB. |
| Female sex associated with longer delays. | 9(20, 35, 36, 44, 51, 64, 65, 139, 144) | Overall, a positive association was found between female sex and increased duration of DD or DUB or rates of DC to BD, although one study showed a longer DC duration for males.(64) |
| **2. Delayed diagnosis – associated outcomes** |  |  |
| **Theme 1: Association of delayed diagnosis with illness course outcomes post-diagnosis** |  |  |
| **Sub-theme** | **Studies including subthemes** | **Key finding** |
| Worsened overall illness course | 8(44, 46, 49, 55, 60, 63, 142, 143) | A positive association was found between increased duration of DD and DUB, and worse clinical outcomes post BD diagnosis, including longer duration of episodes, increased frequency of relapses and lower remission rates, reduced risk assessment and self-management. |
| Increased incidence of suicide attempts. | 9(20, 36, 44, 46, 47, 135, 136, 138, 139) | A positive association was found between increased duration of DD and DUB, and increased incidence of suicide attempts post BD diagnosis |
| Poorer outcomes through anti-depressant monotherapy | 8(20, 39, 44, 47, 57, 72, 136, 147) | A positive association was found between increased duration of DD and DUB and experiencing poorer outcomes due to previous anti-depressant monotherapy post BD-diagnosis. |
| Worsened physical health and general medical outcomes | 2(51, 137) | A positive association was found between increased duration of DD and DUB and experiencing poorer physical health/general medical outcomes post BD-diagnosis. |
| Increased comorbidity | 2(36, 50) | A positive association was found between increased duration of DD and DUB and experiencing increased comorbidity with anxiety disorder or substance abuse post BD-diagnosis. |
| **Theme 2: Association of delayed diagnosis with worsened clinical outcomes post-diagnosis** |  |  |
| Longer or increased hospitalisations. | 6(35, 46, 49, 58, 135, 138) | A positive association was found between increased duration of DD and DUB and experiencing longer or increased frequency of hospitalisations post BD-diagnosis. |
| Delay in providing or accessing any or appropriate treatment/intervention | 5(39, 58, 61, 63, 65) | A positive association was found between increased duration of DD and DUB and experiencing longer or increased delay in providing any or appropriate treatment/intervention post BD-diagnosis, and insufficient information can lead to service user difficulties in making informed treatment decisions. |
| Service user loss of faith and confidence in healthcare system | 1(58) | Data from one of the qualitative studies suggested associations between increased duration of DD and increased service user loss of faith and confidence in healthcare system. |
| Higher costs of care | 1(43) | A positive association was found between increased duration of DD due to misdiagnosis as major depressive disorder and increased cost of care post BD-diagnosis. |
| **Theme 3: Association of delayed diagnosis with worsened social-economic and environmental outcomes post-diagnosis** |  |  |
| Worsened socio-environmental outcomes | 5(49, 60, 63, 137, 141) | A positive association was found between increased duration of DD and DUB and varied worsened socio-environmental outcomes post BD-diagnosis, including increased isolation and unemployment and decreased functioning. |
| **3. Delayed diagnosis – suggested avenues for improvement** |  |  |
| **Theme 1: Clinical care (1) – The need for implementing improved screening processes** |  |  |
| **Sub-theme** | **Studies including subthemes** | **Key finding** |
| Screen for mixed, atypical or rapid-cycling BD in service users presenting with depression. | 9(20, 60, 65, 136, 138, 139, 142, 144, 149) | Multiple studies suggested that duration of DD and DUB could be reduced by introducing routine screening for current or past hypomania, mixed BD symptoms, or rapid-cycling BD in patients presenting with depression. |
| Routine BD screening when diagnosing with depression. | 5(44, 65, 136, 147, 149) | Multiple studies suggested that duration of DD and DUB could be reduced by introducing routine screening for BD or increased longitudinal evaluation in people being diagnosed with major depressive disorder. |
| Routine BD screening in service users with suicide attempts. | 2(20, 139) | Two studies suggested that duration of DD and DUB could be reduced by introducing routine screening for BD in all cases of suicide attempts or in patients who present with a history of suicide attempts. |
| Screen for family history of BD in service users presenting with depression. | 5(20, 63, 65, 136, 144) | Studies suggested routine screening for family history of BD or mania in patients presenting with depression. Multiple studies suggested that duration of DD and DUB could be reduced by introducing routine screening for family history of BD or mania in patients presenting with depression. |
| Screen for mania or hypomania when first presenting with transient psychosis or substance use disorder | 1(148) | One study suggested that duration of DD and DUB could be reduced by introducing routine screening for mania or hypomania in patients presenting with transient psychosis or substance use disorder. |
| Improved differentiation between BD and personality disorders. | 1(11) | One study suggested that duration of DD and DUB could be reduced by improving screening to differentiate between BD and personality disorders. |
| Team-based assessment | 1(59) | One study suggested that team-based assessment, with family member input could help detecting BD. |
| Tracking illness course over time. | 1(59) | Both clinicians and service users suggested that tracking symptoms over time could help in diagnosing BD. |
| **Theme 2: Clinical care (2) – The need for improved services and clinical education** |  |  |
| Improve services: | 5(37, 38, 59, 60, 140) | Four studies suggested that duration of DD and DUB could be reduced by improving clinical services, through changes such as increasing continuity of care, making services more structured, or by interventions designed to facilitate increased attention to patient opinions and experiences, or increased explanation and involvement for family members. Another study singled out the need for increased support, education, and co-management within primary care. |
| Better clinical education around BD | 6(11, 37, 58, 60, 63, 134) | Some studies suggested that duration of DD and DUB could be reduced by improving clinical education around BD. |
| Early intervention, diagnosis etc. | 4(38, 47, 57, 65) | Some studies suggested that duration of DD and DUB could be reduced by increasing provision for early intervention and detection services for BD. |
| Diagnosis disclosure and provision of information | 1(61) | One study highlighted the need for clinicians to disclose BD diagnosis to service users and provide sufficient information. |
| Explore experience and implications of diagnosis with service users and families. | 1(38) | One study suggested the need for clinicians to explore experience and implications of diagnosis with service users and families as a means of facilitating speedier acceptance of BD diagnosis. |
| **Theme 3: The need for social and environmental interventions** |  |  |
| Community psychoeducation and for service users/families could help diagnosis & also reduce stigma | 6(36-38, 57, 58, 152) | Multiple studies suggested that duration of DD and DUB could be reduced by increasing psychoeducation and awareness surrounding BD and decreasing stigma, or that such initiatives might facilitate speedier acceptance of BD diagnosis. |

| **Theme 4: The need for increased and improved research** |  |  |
| --- | --- | --- |
| Improve research on DD and using the knowledge this research generates | 11(11, 20, 35, 36, 46, 47, 57, 63, 64, 140, 141) | Multiple studies suggested that decreasing duration of DD and DUB might be achieved by specific improvements to research on DD and DUB itself, and improving use of the knowledge generated by this research. |
| Increase and improve research to differentiate BD from major depressive disorder. | 2(135, 147) | Two studies suggested that decreasing duration of DD and DUB might be achieved through increasing and improving research on differentiating BD from major depressive disorder. |
| Increase and improve research on specific BD sub-types and symptoms | 2(35, 63) | Two studies suggested that decreasing duration of DD and DUB might be achieved through increasing and improving research on differentiating between different BD subtypes and symptoms. |
| Conduct qualitative work on BD suicide assessment and presentation. | 1(39) | One study suggested that qualitative research may offer insights into clinicians' failure to evaluate risk and poor communication relating to suicide in BD and more 'high-functioning' presentations. |
| Increase/improve research on comorbidities | 2(60, 142) | Two studies suggested that decreasing duration of DD and DUB might be achieved through increasing and improving research on BD comorbidities. |
| Conduct research on the diagnostic process | 2(11, 60) | One study suggested that decreasing duration of DD and DUB might be achieved through research which focuses specifically on the BD diagnostic process, with another study suggesting that there should be research focusing on the role of psychologists in the diagnostic process. |
| Increase and improve research on demographic, socioeconomic and cultural associations | 3(35, 60, 139) | Three studies suggested that decreasing duration of DD and DUB might be achieved through increasing and improving research on demographic, socioeconomic and cultural associations with DD and DUB in BD. |
| Need for common diagnostic markers and improved diagnostic tests/tools | 3(11, 63, 135) | Three studies suggested that decreasing duration of DD and DUB might be achieved through using research to develop improved and more homogeneous diagnostic markers and assessment tools for BD. |

#### 4.1.1 Diverse bipolar presentations

For example, multiple included studies identified higher frequency of hypomanic, mixed, rapid-cycling and atypical depressive presentations as associated precursors. One illustrative example is Abhari et al. 2013, which found that broader diagnostic frameworks incorporating mixed and anti-depressant-induced manic/hypomanic features increased BD diagnosis amongst participants with depression from 18.21% to 53.9%.(72) It seems that an over-reliance on classic binary diagnostic distinctions between manic and depressive states, with insufficient attention to prevalent but often neglected mixed, hypomanic, or rapidly shifting presentations may be exacerbating diagnostic delays.(15, 73) With the exception of one short qualitative synthesis identifying ‘subthreshold manic symptoms’ as a precursor to delayed diagnosis,(18) previous research synthesis is dominated by these classificatory polarities. The need to expand our understanding of depressive, mixed, hypomanic, and rapid-cycling mood states is also supported by extensive research showing limitations in current understanding and consensus on classification and treatment of bipolar depressive states, despite well-documented prevalence and poor outcomes.(74–77) (1, 6, 14, 16, 71, 73, 78–90)

#### 4.1.2 Comorbidities and prior diagnoses

Our mixed findings about association of diagnostic delays with substance abuse and family history were consistent with systematic reviews by Scott at al. 2022 and Keramatian et al. 2025, but diverged from Ratheesh et al. 2017. However, we also identified many other associated conditions not highlighted in previous reviews, including: intellectual disabilities, PTSD, personality disorders, and general health conditions. Our finding that delays were generally shorter in BD with a history of psychotic features is consistent with identification of psychosis as associated with shorter DD in previous reviews. Nevertheless, we also found that psychotic features, particularly associated with hypomanic/manic presentations, can lead to an initial psychotic disorder diagnosis and, thereby, to diagnostic delay, even if eventual delays may average shorter duration than without psychotic features.

#### 4.1.3 Clinical pathways and socio-economic factors

We identified themes and subthemes relating to clinical pathways and care, together with socio-economic, environmental, and demographic factors which augment and sometimes challenge findings from previous systematic reviews. Our review was consistent with previous reviews in identifying potential association between female-sex and increased delays,(17, 91–94) although one included study found that early onset and delayed DD association was stronger for males than females.(64) Unlike Scott et al. 2022, we did not identify the association of diagnostic delays with history of involuntary detention or childhood neglect and had mixed findings for associations with prior hospitalisation (8), due to our exclusion of studies with a paediatric majority population or non-BD-specific focus.

Other themes not identified in previous reviews included mixed and complex associations with socio-economic status. Although lower socio-economic status was generally associated with increased delay duration, we also found some evidence that higher social functioning in BD-I presentations was associated with increased delays, and identified similar complexities relating to social isolation. Inclusion of qualitative studies added numerous insights, including associations between diagnostic delay and factors such as stigma and lack of psychoeducation, or concerns relating to BD and diagnosis, as documented in other qualitative literature on BD.(95–99) Qualitative evidence presented complex pictures of stakeholder reluctance to diagnose and accept BD, which did not necessarily correlate with treatment acceptance, as well as clinical reluctance to diagnose BD in service users with high-functioning presentations.(37, 60)

#### 4.1.4 Outcomes of delayed diagnosis

In general, our review corroborates many suggestions in Scott and Leboyer’s 2011 narrative review of consequences of diagnostic delay, including: increased use of anti-depressant monotherapy, suicide attempts, hospitalisations, illness episodes, poorer socio-economic outcomes and quality of life, and higher costs of care.(17) However, we identified many additional associated outcomes, including poorer physical health outcomes, longer delays in accessing appropriate treatment even post-diagnosis, longer episode duration, poorer risk assessment and self-management. Interestingly, qualitative studies also included findings relating to the effect of diagnostic delays on service users, including reduced faith in healthcare systems and feeling insufficiently informed, which might well affect outcomes.

### 4.2 Avenues for improvement

The most widely suggested avenue for improvement was, understandably, routine screening for BD amongst service users treated for varied presentations of depression or suicidality. However, the need for breadth in future approaches was also evidenced by wide-ranging suggestions about the need for improved clinical care, including clinical education, provision of early intervention services, continuity of care and community psychoeducation. Qualitative studies highlighted collaborative elements of the diagnostic process, including working with service users and their families on team-based assessment, mood tracking, community psychoeducation, diagnostic disclosure and information, and exploring the experience and implications of diagnosis in order to improve BD diagnosis acceptance and reduce stigma. The most common suggestion relating to research was increased research on diagnostic delays and, once again, the breadth of suggestions was notable, including more research on comorbidities, socio-environmental and cultural associations, improved tools and markers for BD assessment and diagnosis, including specific BD-subtypes and symptoms. Qualitative studies again highlighted the collaborative and interpersonal elements of the clinical process, suggesting, for example, further qualitative research exploring weakness in clinical risk assessment, managing suicidality, and identification of BD amongst high-functioning presentations. The urgent need for increased and improved interventions and research relating to DD seems undeniable, with many useful suggestions identified.

### 4.3 Challenges for future research and clinical practice

Before presenting some recommendations for improving the diagnostic process, we must emphasise the need for caution and recognition of related complexities and uncertainties. For example, the current over-reliance on classic binary classificatory distinctions between mania and depression does not reflect the complex range of bipolar presentations and thereby presents potential limitations to implementation of successful BD screening. In general, we lack both consensus about classification and assessment of BD and subtypes, and reliable biomarkers.(15, 100–102) For example, some question the phenomenological validity of the BD-II classification and favour dimensional models,(103) while recent research suggests distinct ‘genetic architectures’ differentiating BD-I and BD-II.(104) Recent developments in genetic, neuroscientific, epidemiological, computational, and multidisciplinary research on differentiating BD from major depression may be useful,(15, 105–112) although applicability remains limited by uncertainties relating to both environmental factors and biomarkers, and considerable overlap of potential predictors with other severe mental illness.(113)

Similar challenges face early BD intervention, a common suggestion in included studies and wider research, which seems particularly important given the association of early age of onset and diagnostic delays.(5, 6, 16, 18, 114–116) However, the heterogeneity and absence of distinct patterns in prodromal BD increases the difficulties of identifying and differentiating early-onset BD from other mood fluctuations associated with adolescence and early adulthood.(117) Early BD presentation often resembles unipolar depression with mixed, fluctuating, or less extreme affective presentations.(1, 118–120) (16, 100) Moreover, while earlier detection could potentially reduce prescription of antidepressant monotherapy, the common depressive presentation of early BD may actually mean that anti-depressant induced polarity switches often constitute the first detectable BD symptom. Potential applicability of evidence-based interventions for early intervention in psychosis(8, 121–123) may also be limited by factors including the more episodic and heterogenous presentation of BD.(124)

We must be cautious about inferring causation from association with particular precursors and framing outcomes as straightforward consequences of DD itself.(16, 17) Our stratification of findings into associated precursors and outcomes indicates complex interrelationships and suggests that earlier diagnosis alone may be insufficient to improve outcomes significantly.(7) For example, while mixed, suicidal, atypical depressive and BD-II presentations were all identified as associated precursors, response rates to available treatment options and general outcomes for these presentations, even when recognised, remain comparatively poor.(6, 125) Additionally, increased incidence of suicide attempts persists even post-diagnosis in those experiencing longer diagnostic delays.

Future research should avoid over-simplification or exaggeration of the standalone benefits of diagnosis. While accurate diagnosis may facilitate timely access to appropriate treatment, reduced antidepressant monotherapy, psychoeducation, guidance for self-management, and other tailored interventions, multiple challenges persist. These include limited understanding of non-classically manic presentations, inadequate resources, education, and suboptimal treatment options for BD management for many individuals and presentations, with high side effect burden and treatment resistance. Conventional clinical guidelines for preventative maintenance treatment with mood-stabilisers have been questioned, due to weak evidence for effectiveness and user desirability.(126) Furthermore, qualitative studies included here highlight reasons for negative views of diagnosis, including stigma, rejection by loved ones,(60) poor BD outcomes, perceiving diagnosis as indeterminate and unscientific,(57, 60) lack of specialist care and insufficient information post-diagnosis.(37, 61)

Finally, estimated delay duration may have limitations concerning reliability and significance. BD diagnosis is not always feasible based on early symptoms,(7, 16) and calculations of diagnostic delay may overlook potentially large numbers of suicide deaths prior to diagnosis, despite associations between suicide attempts and diagnostic delay, general high incidence of suicide attempts in early BD and lethality of BD suicide attempts.(6, 127) Regarding research on “duration of untreated bipolar” (DUB), caution is needed in defining appropriate BD treatments and examining the interrelationship of treatment and diagnosis, given diverse BD presentations, treatment options, and high treatment resistance. While DUB figures are generally based on prescription of mood stabilisers and antipsychotics, the management of bipolar symptoms, regardless of correct diagnosis, may involve a wider range of interventions. These include ECT,(128) benzodiazepines,(129) while less severe symptoms may respond to self-management, or psychosocial interventions.

### 4.4 Study limitations

Our study has several limitations. Key findings are contingent on variables selected in included studies and important issues may therefore have been overlooked. Heterogeneity and potential bias are increased through including small qualitative studies, with limited broad representativeness, while suggested improvements we identified, though interesting, remain subjective. With all studies conducted in high or middle-income countries, findings may not generalise to low or lower-middle income countries, where stigma surrounding severe mental illness and limited specialist healthcare may have distinct effects on diagnosis. Due to the complexities of the search and analysis process, it was not feasible to substantially update searches beyond the database and reference searches completed in July 2024 and January 2025 respectively, although our discussion includes evidence from a 2025 systematic review (16). While we judged it unlikely that any newly published evidence during 2025 would materially alter our conclusions, this remains a limitation. Given our broad inclusion criteria, despite rigorous methods, certain article inclusion or exclusion may be questioned. English-language inclusion restrictions may exclude relevant research. Nevertheless, our use of narrative synthesis, rather than meta-analysis, and relatively consistent findings suggest that these limitations are unlikely to substantially impact on key conclusions.

### 4.5 Implications for future research and practice

While prior reviews suggest that refining DD-related measures to overcome methodological heterogeneity is a future research priority,(7, 8, 16, 18, 19) we suggest that achieving swifter BD diagnosis, with more targeted and effective management, requires new approaches with increased breadth, granularity, and complexity. Most importantly perhaps, the range and diversity of precursors, outcomes, and suggested interventions identified here demonstrate the complex interrelationship of BD diagnosis with other aspects of BD where current limitations in understanding, clinical consensus, and appropriate management are most profound. This suggests that, to improve BD diagnosis, we need to go beyond examining DD in isolation. It also highlights the substantial limitations surrounding the current classification, understanding, and management of BD, compared to other health conditions.

Despite these caveats and limitations, the need to improve BD diagnosis seems incontrovertible, with several key areas identified for future work. These include increasing understanding of: mixed, hypomanic, ‘atypical’, and rapid cycling presentations; the interrelationship of suicidality and diagnosis, given persistence of suicide risk following diagnosis; complex interrelationships between BD and diverse mental and physical health conditions;(130–132) diagnostic processes in lower-and-middle income countries; influence of clinical, socio-economic, cultural, and demographic factors; stakeholder perspectives, including building trust in healthcare systems following diagnostic delays.(16) Importantly, the identification of many distinct themes in qualitative research, particularly relating to collaborative components of diagnostic processes, highlights the need for further qualitative, mixed-methods, and co-produced research. Finally, to progress beyond the current impasse in BD diagnosis towards swifter and more targeted identification and management, it seems we must design future related research and interventions to reflect the complexity of the diagnostic process and its interrelationship with multiple broader challenges associated with management of BD.

## Supporting information

Supplementary Materials Delayed Diagnosis Bipolar Systematic Review

## Data Availability

All data relevant to this review are included in the manuscript

## Declarations Supplementary information

The supplementary information is available online at:

## Author contributions

TG, MA, ST, and TW contributed to the conception and design of this project. TG and MA devised and refined the review and led, both jointly and independently, on all areas of searches, analysis, and data extraction. TW and ST assisted in reviewing the study at all stages, and HA assisted in reviewing initial screening and analysis.

## Data availability statement

Data availability is not applicable to this article as no new data were created or analysed in this study.

