## Supplementary Materials Delayed Diagnosis Bipolar Systematic Review for "A complex interventions approach to delayed diagnosis in bipolar disorder. a co-produced systematic review and narrative synthesis of associated factors"

**Supplementary Information for “The complexity of delayed diagnosis in bipolar disorder. a systematic review and narrative synthesis of associated precursors, outcomes, and suggested avenues for improvement.”**

Gergel, Tania^1,2,3,^ and Al-Janabi, Mariam^1*^., Talwar, Shivangi^1^., Ahmed, Haleemah ^1^., & Wright, Talen^.2^

*Joint first authors

### 1: Note on project development

This project began as a study led by MA and supervised by TG. At the outset, the aim was to explore the phenomenon of delayed diagnosis in bipolar disorder, giving an ‘up-to-date estimation of length of time from onset of symptoms to a diagnosis of bipolar’, with the secondary aims being “Provide further evidence of potential causes and consequences because of delayed diagnosis.” This design can be seen in the initial Prospero (PROSPERO 2024 CRD42024537132) protocol submission Version 1.0. As the team worked on this project, we began to understand that, not only was quantitative synthesis limited by cross-study heterogeneity, but that there was a more urgent need to understand the breadth and complexity of delayed diagnosis in bipolar, in a way which could incorporate the range and diversity of evidence available. Changes to our research aims, design, and methodology are reflected in amendments of our Protocol Version 1, with these developments captured in Versions 2-4, records of which can be found at <https://www.crd.york.ac.uk/PROSPERO/view/CRD42024537132> .

### 2: PRISMA Checklist (with adaptations for narrative synthesis).

| **Section and Topic** | **Item #** | **Checklist item** | **Location where item is reported** |
| --- | --- | --- | --- |
| **TITLE** | | |  |
| Title | 1 | Identify the report as a systematic review. | 1 |
| **ABSTRACT** | | |  |
| Abstract | 2 | See the PRISMA 2020 for Abstracts checklist. | 2 |
| **INTRODUCTION** | | |  |
| Rationale | 3 | Describe the rationale for the review in the context of existing knowledge. | 3 |
| Objectives | 4 | Provide an explicit statement of the objective(s) or question(s) the review addresses. | 3-4 |
| **METHODS** | | |  |
| Eligibility criteria | 5 | Specify the inclusion and exclusion criteria for the review and how studies were grouped for the syntheses. | 4 |
| Information sources | 6 | Specify all databases, registers, websites, organisations, reference lists and other sources searched or consulted to identify studies. Specify the date when each source was last searched or consulted. | 3 and Supplementary Materials 3 |
| Search strategy | 7 | Present the full search strategies for all databases, registers and websites, including any filters and limits used. | 3 and Appendix 3 |
| Selection process | 8 | Specify the methods used to decide whether a study met the inclusion criteria of the review, including how many reviewers screened each record and each report retrieved, whether they worked independently, and if applicable, details of automation tools used in the process. | 3 |
| Data collection process | 9 | Specify the methods used to collect data from reports, including how many reviewers collected data from each report, whether they worked independently, any processes for obtaining or confirming data from study investigators, and if applicable, details of automation tools used in the process. | 3-4 |
| Data items | 10a | List and define all outcomes for which data were sought. Specify whether all results that were compatible with each outcome domain in each study were sought (e.g. for all measures, time points, analyses), and if not, the methods used to decide which results to collect. | 3-4 |
|  | 10b | List and define all other variables for which data were sought (e.g. participant and intervention characteristics, funding sources). Describe any assumptions made about any missing or unclear information. | 3-4 |
| Study risk of bias assessment | 11 | Specify the methods used to assess risk of bias in the included studies, including details of the tool(s) used, how many reviewers assessed each study and whether they worked independently, and if applicable, details of automation tools used in the process. | 4 |
| Effect measures | 12 | Specify for each outcome the effect measure(s) (e.g. risk ratio, mean difference) used in the synthesis or presentation of results. | N/A |
| Synthesis methods | 13a | Describe the processes used to decide which studies were eligible for each synthesis (e.g. tabulating the study intervention characteristics and comparing against the planned groups for each synthesis (item #5)). Methods for narrative synthesis are described in detail in data synthesis section of the Methods section. | 5 |
|  | 13b | Describe any methods required to prepare the data for presentation or synthesis, such as handling of missing summary statistics, or data conversions. | 5 |
|  | 13c | Describe any methods used to tabulate or visually display results of individual studies and syntheses. | 5 |
|  | 13d | Describe any methods used to synthesize results and provide a rationale for the choice(s). If meta-analysis was performed, describe the model(s), method(s) to identify the presence and extent of statistical heterogeneity, and software package(s) used. | 5 and Supplementary Materials Section 7. |
|  | 13e | Describe any methods used to explore possible causes of heterogeneity among study results (e.g. subgroup analysis, meta-regression). | N/A |
|  | 13f | Describe any sensitivity analyses conducted to assess robustness of the synthesized results. | N/A |
| Reporting bias assessment | 14 | Describe any methods used to assess risk of bias due to missing results in a synthesis (arising from reporting biases). | N/A |
| Certainty assessment | 15 | Describe any methods used to assess certainty (or confidence) in the body of evidence for an outcome. | N/A |
| **RESULTS** | | |  |
| Study selection | 16a | Describe the results of the search and selection process, from the number of records identified in the search to the number of studies included in the review, ideally using a flow diagram. | 5 and Figure 1 |
|  | 16b | Cite studies that might appear to meet the inclusion criteria, but which were excluded, and explain why they were excluded. | Figure 1 and Supplementary Materials Section 4 |
| Study characteristics | 17 | Cite each included study and present its characteristics. | Table 2: Study characteristics. |
| Risk of bias in studies | 18 | Present assessments of risk of bias for each included study. | 5 and Supplementary Materials Section 5. |
| Results of individual studies | 19 | For all outcomes, present, for each study: (a) summary statistics for each group (where appropriate) and (b) an effect estimate and its precision (e.g. confidence/credible interval), ideally using structured tables or plots. | N/A |
| Results of syntheses | 20a | For each synthesis, briefly summarise the characteristics among contributing studies. | 5-6 |
|  | 20b | Present results of all statistical syntheses conducted. If meta-analysis was done, present for each the summary estimate and its precision (e.g. confidence/credible interval) and measures of statistical heterogeneity. If comparing groups, describe the direction of the effect. | Qualitative synthesis of themes presented in Table 3. |
|  | 20c | Present results of all investigations of possible causes of heterogeneity among study results. | Supplementary Materials Section 6. |
|  | 20d | Present results of all sensitivity analyses conducted to assess the robustness of the synthesized results. | N/A |
| Reporting biases | 21 | Present assessments of risk of bias due to missing results (arising from reporting biases) for each synthesis assessed. | N/A |
| Certainty of evidence | 22 | Present assessments of certainty (or confidence) in the body of evidence for each outcome assessed. | N/A |
| **DISCUSSION** | | |  |
| Discussion | 23a | Provide a general interpretation of the results in the context of other evidence. | 8-9 |
|  | 23b | Discuss any limitations of the evidence included in the review. | 10-11 |
|  | 23c | Discuss any limitations of the review processes used. | 11-12 |
|  | 23d | Discuss implications of the results for practice, policy, and future research. | 12 |
| **OTHER INFORMATION** | | |  |
| Registration and protocol | 24a | Provide registration information for the review, including register name and registration number, or state that the review was not registered. | 4 |
|  | 24b | Indicate where the review protocol can be accessed, or state that a protocol was not prepared. | 4 |
|  | 24c | Describe and explain any amendments to information provided at registration or in the protocol. | Supplementary Materials Section 1. |
| Support | 25 | Describe sources of financial or non-financial support for the review, and the role of the funders or sponsors in the review. | 13 |
| Competing interests | 26 | Declare any competing interests of review authors. | 13 |
| Availability of data, code and other materials | 27 | Report which of the following are publicly available and where they can be found: template data collection forms; data extracted from included studies; data used for all analyses; analytic code; any other materials used in the review. | Table 2 and Table 3 |

### 3: Full search strategies

**Literature search: Delayed diagnosis and treatment of bipolar disorder and diagnostic conversion to bipolar disorder.**

Key search terms included the main constructs and their derivatives, designed to capture core concepts of delay, diagnosis and/or treatment, and BD: delay or “delay*” or “missed” or “unrecognised” or “unrecognized” or “unrecogni*” or “untreated” or “untreat*” or misdiagnos* or “misdiagnosis” or “misdiagnoz*” or diagnos* or “diagnosis” or “diagnoz*” AND bipolar disorder or “bipolar affective disorder” or “bipolar*” or “manic depress*” or “manic depression” or “mani*” or “bipolar NOS”.

Date of search: 15^th^ July 2024

- MEDLINE (OVID), n=4989
- PsychINFO (OVID), n=4663
- Embase (OVID), n=4122
- Web of Science, n=6754

Total= 20528

Duplicates excluded=9282

De-duplicated, to screen, n=11246

**Search Strategies**

1. **Ovid MEDLINE(R) ALL <1946 to July 15, 2024>**

| 1 | (psychiatry or mental disorders or psychology).sh. | 239,202 |
| --- | --- | --- |
| 2 | ("mental disorder*" or "mental illness*" or psychiatr*).tw. | 359,329 |
| 3 | 1 or 2 | 482,551 |
| 4 | Bipolar disorder.sh. | 46,600 |
| 5 | ("bipolar disorder*" or "bipolar affective disorder*" or bipolar* or "manic depressi*" or manic or mania or "bipolar NOS").tw. | 89,067 |
| 6 | 4 or 5 | 100,400 |
| 7 | 3 and 6 | 23,817 |
| 8 | (Diagnostic errors or Delayed Diagnosis).sh. | 48,558 |
| 9 | ("unrecognised" or "unrecognized" or "unrecogni*" or "untreated" or "untreat*" or misdiagnos* or "misdiagnosis" or "misdiagnoz*" or diagnos* or "diagnosis" or "diagnoz*").tw. | 3,467,963 |
| 10 | 8 or 9 | 3,486,173 |
| 11 | 7 and 10 | 9,807 |
| 12 | limit 11 to (english language and humans and "all adult (19 plus years)") | 4,989 |

2. **APA PsycInfo <1806 to July 2024 Week 3>**

| 1 | (Bipolar Disorder or Cyclothymic Disorder or Mania or Bipolar I Disorder or Bipolar II Disorder).mp. | 52,919 |
| --- | --- | --- |
| 2 | ("bipolar disorder*" or "bipolar affective disorder*" or bipolar* or "manic depressi*" or manic or mania or "bipolar NOS").tw. | 59,256 |
| 3 | 1 or 2 | 64,295 |
| 4 | (Misdiagnosis or Treatment Delay or Diagnostic Errors).mp. | 4,090 |
| 5 | ("unrecognised" or "unrecognized" or "unrecogni*" or "untreated" or misdiagnos* or "misdiagnosis" or "misdiagnoz*" or diagnos* or "diagnosis" or "diagnoz*").tw. | 400,727 |
| 6 | 4 or 5 | 401,134 |
| 7 | 3 and 6 | 20,032 |
| 8 | limit 7 to (peer reviewed journal and human and english language and "300 adulthood " and "remove medline records") | 4,663 |

3. **Embase Classic+Embase <1947 to 2024 July 15>**

| 1 | *psychiatry/ | 57,937 |
| --- | --- | --- |
| 2 | *mental disease/ | 137,156 |
| 3 | ("mental disorder*" or "mental illness*" or psychiatr*).tw. | 521,486 |
| 4 | 1 or 2 or 3 | 603,740 |
| 5 | *bipolar disorder/ | 34,244 |
| 6 | ("bipolar disorder*" or "bipolar affective disorder*" or bipolar* or "manic depressi*" or manic or mania or "bipolar NOS").tw. | 131,798 |
| 7 | 5 or 6 | 134,768 |
| 8 | *diagnostic error/ | 12,251 |
| 9 | ("unrecognised" or "unrecognized" or "unrecogni*" or "untreated" or misdiagnos* or "misdiagnosis" or "misdiagnoz*" or diagnos* or "diagnosis" or "diagnoz*").tw. | 5,285,139 |
| 10 | 4 and 7 | 35,847 |
| 11 | 8 or 9 | 5,288,005 |
| 12 | 10 and 11 | 16,751 |
| 13 | limit 12 to (human and english language and "remove medline records" and adult <18 to 64 years>) | 4,122 |

4. **Web of Science – Date run: Mon Jul 15 2024 10:35:39 GMT+**

| 1 | (ALL=((psychiatry or mental disorders or psychology).KP)) OR TS=(("mental disorder*" or "mental illness*" or psychiatr*)) | 469741 |
| --- | --- | --- |
| 2 | (ALL=(Bipolar disorder.KP)) OR TS=(("bipolar disorder*" or "bipolar affective disorder*" or bipolar* or "manic depressi*" or manic or mania or "bipolar NOS").) | 195178 |
| 3 | #2 AND #1 | 29052 |
| 4 | (ALL=((Diagnostic errors or Delayed Diagnosis).KP)) OR TS=(("unrecognised" or "unrecognized" or "unrecogni*" or "untreated" or "untreat*" or misdiagnos* or "misdiagnosis" or "misdiagnoz*" or diagnos* or "diagnosis" or "diagnoz*").) | 4075252 |
| 5 | #4 AND #3 | 12223 |
| 6 | Search: #4 AND #3 and English (Languages) and 1.21 Psychiatry or 6.24 Psychiatry & Psychology (Citation Topics Meso) | 6754 |

### 4: Examples of excluded studies

**Studies not insufficient focus on BD or where BD-specific data difficult to extract:**

Berk M, Dodd S, Callaly P, Berk L, Fitzgerald P, de Castella AR, Filia S, Filia K, Tahtalian S, Biffin F, Kelin K, Smith M, Montgomery W, Kulkarni J. History of illness prior to a diagnosis of bipolar disorder or schizoaffective disorder. J Affect Disord. 2007 Nov;103(1-3):181-6. doi: 10.1016/j.jad.2007.01.027. Epub 2007 Feb 26. PMID: 17324469.

Chen YR, Swann AC, Johnson BA. Stability of diagnosis in bipolar disorder. J Nerv Ment Dis. 1998 Jan;186(1):17-23. doi: 10.1097/00005053-199801000-00004. PMID: 9457143.

Nguyen T, Tran T, Green S, Hsueh A, Tran T, Tran H, Fisher J. Delays to diagnosis among people with severe mental illness in rural Vietnam, a population-based cross-sectional survey. BMC Psychiatry. 2019 Dec 4;19(1):385. doi: 10.1186/s12888-019-2367-1. PMID: 31801486; PMCID: PMC6894253.

Okan Ibiloglu A, Caykoylu A. The comorbidity of anxiety disorders in bipolar I and bipolar II patients among Turkish population. J Anxiety Disord. 2011 Jun;25(5):661-7. doi: 10.1016/j.janxdis.2011.02.008. Epub 2011 Feb 21. PMID: 21411273.

**Insufficient focus on delayed bipolar diagnosis:**

Benazzi F. Clinical differences between bipolar II depression and unipolar major depressive disorder: lack of an effect of age. J Affect Disord. 2003 Jul;75(2):191-5. doi: 10.1016/s0165-0327(02)00047-2. PMID: 12798259.

Mianji F, Kirmayer LJ. Help-seeking strategies and treatment experiences among individuals diagnosed with Bipolar Spectrum Disorder in Iran: A qualitative study. Transcult Psychiatry. 2023 Apr;60(2):201-214. doi: 10.1177/13634615221127855. Epub 2022 Oct 17. PMID: 36245238; PMCID: PMC10150414.

García-López A, Ezquiaga E, De Dios C, Agud JL. Depressive symptoms in early- and late-onset older bipolar patients compared with younger ones. Int J Geriatr Psychiatry. 2017 Feb;32(2):201-207. doi: 10.1002/gps.4465. Epub 2016 Mar 27. PMID: 27017999.

Inoue T, Inagaki Y, Kimura T, Shirakawa O. Prevalence and predictors of bipolar disorders in patients with a major depressive episode: the Japanese epidemiological trial with latest measure of bipolar disorder (JET-LMBP). J Affect Disord. 2015 Mar 15;174:535-41. doi: 10.1016/j.jad.2014.12.023. Epub 2014 Dec 15. PMID: 25556671.

Morgan C, Ashcroft DM, Chew-Graham CA, Sperrin M, Webb RT, Francis A, Scott J, Yung AR. Identifying prior signals of bipolar disorder using primary care electronic health records: a nested case-control study. Br J Gen Pract. 2024 Feb 29;74(740):e165-e173. doi: 10.3399/BJGP.2022.0286. PMID: 38325893; PMCID: PMC10877620.

Stewart C, El-Mallakh RS. Is bipolar disorder overdiagnosed among patients with substance abuse? Bipolar Disord. 2007 Sep;9(6):646-8. doi: 10.1111/j.1399-5618.2007.00465.x. PMID: 17845280

Bukh JD, Andersen PK, Kessing LV. Rates and predictors of remission, recurrence and conversion to bipolar disorder after the first lifetime episode of depression--a prospective 5-year follow-up study. Psychol Med. 2016 Apr;46(6):1151-61. doi: 10.1017/S0033291715002676. Epub 2016 Jan 8. PMID: 26743873.

**Predominant focus on effectiveness of intervention (e.g. screening and prediction):**

Agius M, Murphy CL, Zaman R. Under-diagnosis of bipolar affective disorder in A bedford CMHT. Psychiatr Danub. 2010 Nov;22 Suppl 1:S36-7. PMID: 21057399.

Smith DJ, Griffiths E, Kelly M, Hood K, Craddock N, Simpson SA. Unrecognised bipolar disorder in primary care patients with depression. Br J Psychiatry. 2011 Jul;199(1):49-56. doi: 10.1192/bjp.bp.110.083840. Epub 2011 Feb 3. PMID: 21292927.

Das AK, Olfson M, Gameroff MJ, Pilowsky DJ, Blanco C, Feder A, Gross R, Neria Y, Lantigua R, Shea S, Weissman MM. Screening for bipolar disorder in a primary care practice. JAMA. 2005 Feb 23;293(8):956-63. doi: 10.1001/jama.293.8.956. PMID: 15728166.

Benacek J, Martin-Key NA, Spadaro B, Tomasik J, Bahn S. Using decision-analysis modelling to estimate the economic impact of the identification of unrecognised bipolar disorder in primary care: the untapped potential of screening. Int J Bipolar Disord. 2022 Jun 10;10(1):15. doi: 10.1186/s40345-022-00261-9. PMID: 35680705; PMCID: PMC9184689.

Gilman SE, Dupuy JM, Perlis RH. Risks for the transition from major depressive disorder to bipolar disorder in the National Epidemiologic Survey on Alcohol and Related Conditions. J Clin Psychiatry. 2012 Jun;73(6):829-36. doi: 10.4088/JCP.11m06912. Epub 2012 Feb 21. PMID: 22394428; PMCID: PMC3703739.

Takeshima M, Kitamura T, Kitamura M, Kidani T, Tochimoto S, Muramori F, Kosaka K, Hasegawa M, Ueno K, Hamahara S, Kurata K. Impact of depressive mixed state in an emergency psychiatry setting: a marker of bipolar disorder and a possible risk factor for emergency hospitalization. J Affect Disord. 2008 Nov;111(1):52-60. doi: 10.1016/j.jad.2008.02.009. Epub 2008 Mar 19. PMID: 18355924.

**Focus on paediatric BD**

Goetz M, Novak T, Vesela M, Hlavka Z, Brunovsky M, Povazan M, Ptacek R, Sebela A. Early stages of pediatric bipolar disorder: retrospective analysis of a Czech inpatient sample. Neuropsychiatr Dis Treat. 2015 Nov 4;11:2855-64. doi: 10.2147/NDT.S79586. PMID: 26604770; PMCID: PMC4639550.

### 5: Quality Assessment Scores

High ratings were largely due to consistent use of reliable measures, diagnostic categories, and robust methodologies within individual studies. The main reason for deductions in quality ratings was due to a few studies lacking adequate detail of control of confounding variables, analytic detail of the methods, the sample size and selection process in some studies were not justified and could be argued as unrepresentative/ biased. The assessment tool used throughout was the Kmet Scale: Checklist for assessing the Quality of quantitative studies, a tool which is well established for appraising and synthesising quality assessment across studies with diverse methodologies.

| **Author** | **Quality Assessment** | |
| --- | --- | --- |
| Ahmed et al., 2021 | 86% | High Quality |
| Altamura et al., 2010 | 95% | Very High Quality |
| Altamura et al., 2015 | 90% | High Quality |
| Altamura et al., 2018 | 86% | High Quality |
| Azorin et al., 2015 | 95% | Very High Quality |
| Carlborg et al., 2015 | 100% | Very High Quality |
| Drancourt et al., 2013 | 86% | High Quality |
| Isogaya et al., 2021 | 73% | Medium Quality |
| Oyffe et al., 2015 | 77% | Medium Quality |
| Medeiros et al., 2015 | 86% | High Quality |
| Szmulewicz et al., 2019 | 68% | Low Quality |
| Undurraga et al., 2012 | 90% | High Quality |
| Buoli et al., 2021 | 100% | Very High Quality |
| Cha et al., 2009 | 90% | High Quality |
| Gazelle et al 2005 | 90% | High Quality |
| Goldberg et al., 2002 | 95% | Very High Quality |
| Hong et al., 2016 | 95% | Very High Quality |
| Keramatian et al., 2022 | 90% | High Quality |
| Kvitland et al., 2016 | 90% | High Quality |
| Lubloy et al., 2019 | 100% | Very High Quality |
| Maina et al., 2013 | 90% | High Quality |
| Mantere et al., 2008 | 100% | Very High Quality |
| McCraw et al., 2014 | 95% | Very High Quality |
| Murru et al.. 2019 | 90% | High Quality |
| Nery-Fernandes et al., 2012 | 100% | Very High Quality |
| Rosa et al., 2007 | 90% | High Quality |
| Zhang et al., 2017 | 95% | Very High Quality |
| Abhari et al., 2013 | 95% | Very High Quality |
| Angst et al., 2005 | 77% | Medium Quality |
| Bouchra et al., 2017 | 81% | High Quality |
| Chauhan et al., 2022 | 77% | Medium Quality |
| Fritz et al., 2017 | 77% | Medium Quality |
| Hinphet et al., 2021 | 95% | Very High Quality |
| Hughes et al., 2016 | 81% | High Quality |
| Inoue et al., 2006 | 73% | Medium Quality |
| Jo et al ., 2022 | 100% | Very High Quality |
| Kessing., 2005 | 82% | High Quality |
| Kucukalic et al., 2012 | 86% | High Quality |
| McCombs et al., 2007 | 95% | Very High Quality |
| Highet et al., 2004 | 75% | Medium Quality |
| Cerimele et al., 2019 | 85% | High Quality |
| Clements et al., 2019 | 80% | High Quality |
| Delmas et al., 2011 | 65% | Low Quality |
| Johansson & Werbart., 2020 | 90% | High Quality |
| Kashani et al., 2020 | 70% | Medium Quality |
| Maassen et al, 2018 | 90% | High Quality |
| Maassen et al., 2018 | 85% | High Quality |
| Maassen et al., 2019 | 75% | Medium Quality |
| Siegel-Ramsay et al., 2023 | 90% | Very High Quality |

### 6: Additional qualitative synthesis of results relating to measures, delay duration, and diagnostic conversion rates.

DUB: duration of untreated bipolar disorder:

The most commonly examined and consistently defined and measured latency was DUB, which featured in 17 studies. 13 studies gave average DUB (in years), with ten finding an average DUB of between 5.16 years and 10.4 years,(1-9) two reporting a far lower one a far higher DUB.(10-12) Two studies presented differing durations for study subgroups.(9, 13) DUB was generally defined as the time from the onset of illness to initiation of appropriate treatment. Appropriate treatment was specified as mood-stabilising medication in 13 studies, with the definition of initiation varying between the start of the course and the date at which a patient had been on the prescribed medication for an adequate treatment period. Two studies defined appropriate treatment broadly as correct or effective treatment.(10, 11) Finally, two studies simply defined the initiation of treatment as first contact with psychiatric care.(13, 14) While the majority of studies classified onset as the first affective episode, there was a lack of consistency and precision in outlining what constituted an initial affective episode and how this was recorded.

DD: duration of delayed diagnosis.

14 studies focused on DD, with 10 giving average DD (in years). Eight studies gave average DD of between 5.3 and 8 years,(9, 15-22) with two studies reporting far lower and one a far higher DD.(23-25) Five studies presented differing durations for study subgroups.(9, 16, 20, 22, 25) There was substantial heterogeneity in how the DD latency was defined, with nine studies defining this as length of time between the onset of symptoms or mood episode and BD diagnosis, and four as length of time from clinical contact or first psychiatric diagnosis to BD diagnosis.

DC/UBD: rate of diagnostic conversion from depressive disorder diagnosis to bipolar disorder diagnosis or unrecognised bipolar disorder amongst those diagnosed with depression.

12 studies examined unrecognised BD and diagnostic conversion (DC) from depression to BD as the primary exposure with the main measurement being percentage of participants with a major or recurrent depressive disorder or episode who met the diagnostic criteria for BD, but had not received a prior diagnosis of BD. Eight studies found rates of unrecognised BD amongst people diagnosed with depression ranging between 18.9% to 39.84%,(19, 26-32) while one study gave a rate of either 18.21% or 53.9%, depending on whether the DSM-IV-TR or broader Bipolar Specifier criteria were used.(33) Finally, there were three studies with lower rates, ranging from 6.5% to 7.3%.(34-36)

Features of studies with unusually low or high average DD, DUB, or DC.

Any averages for DD or DUB between 5.16 and 5.96 were the duration of lower comparative population subgroups.^48,56,64^ In four studies with much lower reported DUB/DD, study populations were either wholly or predominantly diagnosed with BD-I and/or mania and current or recent inpatients,^38,44,65,66^ while populations in the three studies reporting far lower rates of DC included or were exclusively primary care populations,^40,67^ or exclusively people currently taking antidepressants without accounting for those who had terminated antidepressants due to polarity switch.^41^ The two studies which reported much higher DD/DUB had majority BD-II populations or general medical comorbidities,^51,68^ while one DC study reported a much higher rate if a broader bipolar specifier was used instead of DSM-IV-TR.^69^

### 7: Additional information concerning the use of qualitative narrative synthesis methodology for this systematic review

Some articles on systematic and scoping reviews suggest that qualitative synthesis is more suited to scoping reviews and that reviews which are not primarily focussed on some type of quantitative synthesis, both in relation to results and in assessing risk of bias should be viewed as scoping rather than systematic reviews, particularly those based primarily in physical healthcare, such as Smith and Duncan 2022 “Systematic and scoping reviews: A comparison and overview” DOI: 10.1053/j.semvascsurg.2022.09.001, from *'Seminars in Vascular Surgery'*.

However, particularly in mental health, where there is a lack of distinct biomarkers and a lack of consensus over e.g. diagnostic classifications, methods, and measurements, and where much evidence comes from sources which contain far higher levels of heterogeneity and subjectivity than many areas of physical healthcare, there is strong precedent for using complex and systematic qualitative/narrative synthesis as the basis for systematic reviews. The methods used within this paper are recognised as valid methods for systematic reviews in relation to complex interventions, breadth of aims and factors for consideration, and mixed quant and qual evidence sources for synthesis, for example in Noyes et al. 2018, “Synthesising quantitative and qualitative evidence to inform guidelines on complex interventions: clarifying the purposes, designs and outlining some methods”  DOI: 10.1136/bmjgh-2018-000893, which is given as the primary methodological reference in our paper. Another methodological paper which is useful and also referenced is a chapter by Popay et al 2006: ''Guidance on the conduct of narrative synthesis in systematic reviews: A product from the ESRC Methods Programme" DOI 10.13140/2.1.1018.4643. Further factors and references relating to the validity of using a systematic review rather than scoping review for the current study are explained in the table below (7.1).

Finally, some papers which provide a precedent for the exclusive use of qualitative narrative synthesis for synthesising quantitative and qualitative literature in systematic reviews, and for breadth and complexity of scope, include the following: Uddin et al. 2023 (PMID: 37772412); Gronholm et al. 2017 (PMID: 28196549); Baldwin et al 2025 (PMID: 40717551); Nimmo-Smith et al 2020 (PMID: 32036811); Maguire et al. (PMID: 38600985); Harvey et al. (PMID: 37779607)

#### 7.1 Some factors underpinning choice of systematic review rather than scoping review for the current study/evidence synthesis.

| **Factor** | **Systematic Review** | **Scoping Reviews** | **Our approach** | **Source** |
| --- | --- | --- | --- | --- |
| Key indications re purpose and scope of review | 1. Uncover the international evidence 2. Confirm current practice/ address any variation/ identify new practices 3. Identify and inform areas for future research 4. Identify and investigate conflicting results 5. Produce statements to guide decision-making | Possibly:   1. Precursor to systematic review. 2. Less clarity on 'population, concept, context'. | We fulfilled all 5 key indicators for systematic reviews as identified by Munn.    We clearly identified target population: studies focused on adults with BD.    We clearly identified core 'concept', in line with many a strong body of research studies and other evidence synthesis in this field: i.e. delayed diagnosis and treatment of BD, including failure to recognise BD, in cases where a diagnosis of BD would be indicated clinically.    In addition, we carried out a scoping stage prior to finalising our protocol for a systematic review. | Munn et al. 2018. [Systematic review or scoping review? Guidance for authors when choosing between a systematic or scoping review approach - PMC](https://pmc.ncbi.nlm.nih.gov/articles/PMC6245623/) |
| Aims: | To answer a specific clinical question (hypothesis testing) | To summarise types and quality of literature on a topic, clarify concepts, uncover knowledge gaps | Distinct aims:   - To answer the question: what factors have been identified in research as precursors, outcomes, and potential avenues for improvement relating to delayed diagnosis. - To use complex intervention framework for DD evidence synthesis. - To identify the range of factors relating to diagnostic delay, stratified into associated precursors, outcomes, and avenues for improvement.   Despite the focus on breadth, this does not detract from the systematic nature of the investigation or from the distinct question which is being asked. | Munn et al. 2018 |
| Search methods | A reproducible search must be conducted in at least two (some say four) databases | Specifically defined ahead of time, more likely to use nonindexed sources | 4 databases searched separately with all details of search provided in Appendix, so that search can be reproduced. | Smith and Duncan 2022 [Systematic and scoping reviews: A comparison and overview - ScienceDirect](https://www.sciencedirect.com/science/article/pii/S0895796722000631) |
| Synthesis of findings from individual studies and the generation of ‘summary’ findings^c^ | Yes | No | Synthesis of findings was carried out via a robust framework for qualitative synthesis in systematic reviews, detailed in methodologies, provided in results.    'Summary' findings were generated. | Munn et al. 2018.    For methods used in this paper and their validity as frameworks for systematic reviews in relation to complex interventions and mixtures of quantitative and qualitative evidence for synthesis, please see Noyes et al 2019, referenced in our paper: [Synthesising quantitative and qualitative evidence to inform guidelines on complex interventions: clarifying the purposes, designs and outlining some methods - PMC](https://pmc.ncbi.nlm.nih.gov/articles/PMC6350750/) |
| Prospero registration and A priori review protocol, including Inclusion criteria for topics and study types. | Registered with Prospero | Not registered with Prospero.    A prior protocol is optional.    Protocol may be revised as research explored | Registered with Prospero, listed in article and Appendix, with any amendments to original protocol listed/referenced in Appendix 1 and changed prior to agreeing final protocol and carrying out searches.    During earlier scoping stages of project we refined search terms and study types, but in final study plan and protocol, the study types, topics, and terms were set | Smith and Duncan 2022    Munn et al. 2018. |

21.	Lubloy A, Lilla KJ, Attila N, Peter M. Exploring factors of diagnostic delay for patients with bipolar disorder: A population based cohort study. Journal of Mental Health Policy and Economics. 2019;22(SUPPL 1):S21-S2.
